# Longitudinal home self-collection of capillary blood using *home*RNA correlates interferon and innate viral defense pathways with SARS-CoV-2 viral clearance

**DOI:** 10.1101/2023.01.24.23284913

**Authors:** Fang Yun Lim, Soo-Young Kim, Karisma N. Kulkarni, Rachel L. Blazevic, Louise E. Kimball, Hannah G. Lea, Amanda J. Haack, Maia. S. Gower, Terry Stevens-Ayers, Lea M. Starita, Michael Boeckh, Joshua T. Schiffer, Ollivier Hyrien, Ashleigh B. Theberge, Alpana Waghmare

**Author notes:** Corresponding authors: Ashleigh B. Theberge; Alpana Waghmare.

## Abstract

Blood transcriptional profiling is a powerful tool to evaluate immune responses to infection; however, blood collection via traditional phlebotomy remains a barrier to precise characterization of the immune response in dynamic infections (e.g., respiratory viruses). Here we present an at-home self-collection methodology, *home*RNA, to study the host transcriptional response during acute SARS-CoV-2 infections. This method uniquely enables high frequency measurement of the host immune kinetics in non-hospitalized adults during the acute and most dynamic stage of their infection. COVID-19+ and healthy participants self-collected blood every other day for two weeks with daily nasal swabs and symptom surveys to track viral load kinetics and symptom burden, respectively. While healthy uninfected participants showed remarkably stable immune kinetics with no significant dynamic genes, COVID-19+ participants, on the contrary, depicted a robust response with over 418 dynamic genes associated with interferon and innate viral defense pathways. When stratified by vaccination status, we detected distinct response signatures between unvaccinated and breakthrough (vaccinated) infection subgroups; unvaccinated individuals portrayed a response repertoire characterized by higher innate antiviral responses, interferon signaling, and cytotoxic lymphocyte responses while breakthrough infections portrayed lower levels of interferon signaling and enhanced early cell-mediated response. Leveraging cross-platform longitudinal sampling (nasal swabs and blood), we observed that *IFI27*, a key viral response gene, tracked closely with SARS-CoV-2 viral clearance in individual participants. Taken together, these results demonstrate that at-home sampling can capture key host antiviral responses and facilitate frequent longitudinal sampling to detect transient host immune kinetics during dynamic immune states.

**One Sentence Summary:** Self-blood collection using *home*RNA captures temporal dynamics in host transcriptional immune response during acute SARS-CoV-2 infection.

## INTRODUCTION

The blood transcriptome can offer valuable mechanistic insights into an individual’s immune response to chronic and infectious diseases (*1-3*). In efforts towards personalized medicine, high-dimensional molecular diagnostics based on blood transcriptional signatures have been developed to detect infectious diseases, inform prognosis, and tailor treatment strategies in various cancers and autoimmune diseases (*4-9*). However, these multivariate molecular signature assays remain heavily reliant on traditional phlebotomy, a procedure that is resource-extensive, can be burdensome for patients, and is particularly difficult to perform daily (*10*). Consequently, these limitations restrict access to specimen collection, especially when early and frequent timepoints are desired. Instead of using disease- or treatment-relevant time points (precision sampling), clinical studies often rely on scheduling convenience or clinical staff availability (convenience sampling). In the era of precision medicine, convenience sampling can compromise the predictive values of biomarkers in both clinical testing and during biomarker development studies. Within the context of respiratory viral infection and the rapidly evolving and potentially transient nature of early infection immune kinetics, single timepoint participant samples (cross-sectional/non-longitudinal studies) can be misleading when confounded by each individual’s infection timeline. A home-use device that decentralizes specimen collection while preserving transcriptomic signatures during specimen transport back to a central lab has tremendous potential to catalyze development and application of RNA-based diagnostics and clinical research into the fundamental mechanisms of disease.

Our team recently developed a home-use device (*home*RNA) that enables self-collection of liquid capillary blood coupled with in-field stabilization of blood cellular RNA (*11*). This platform enables larger blood volumes and a potentially less painful user experience compared to prior systems that utilize finger sticks; further, by using a larger volume liquid sample, *home*RNA overcomes many of the limitations of using dried blood spots for transcriptomics. The *home*RNA blood collection kit consists of two major components: i) a Tasso-SST blood collection device (Tasso, Inc.) and ii) a stabilizer tube with embedded fluidics containing RNA stabilizer (**Fig. 1**). We previously assessed its usability in an unsupervised setting using a cohort of healthy volunteers and demonstrated that at-home participant-collected and stabilized blood samples obtained from healthy adult volunteers yielded high RNA quality and sufficient RNA quantity suitable for downstream transcriptomics analysis (*11*). By recruiting study participants across the U.S., we demonstrated the feasibility of decentralized liquid biopsy for transcriptomics-based applications. In clinical study applications, this virtual self-collection strategy makes *home*RNA scalable to geographically disparate multi-site studies over longer study durations while still maintaining i) a systematic collection schedule and standardization of collection protocols across the study and ii) high resolution time-course profiling of the blood transcriptional response across disease- or treatment-relevant time points.

**Figure 1.**
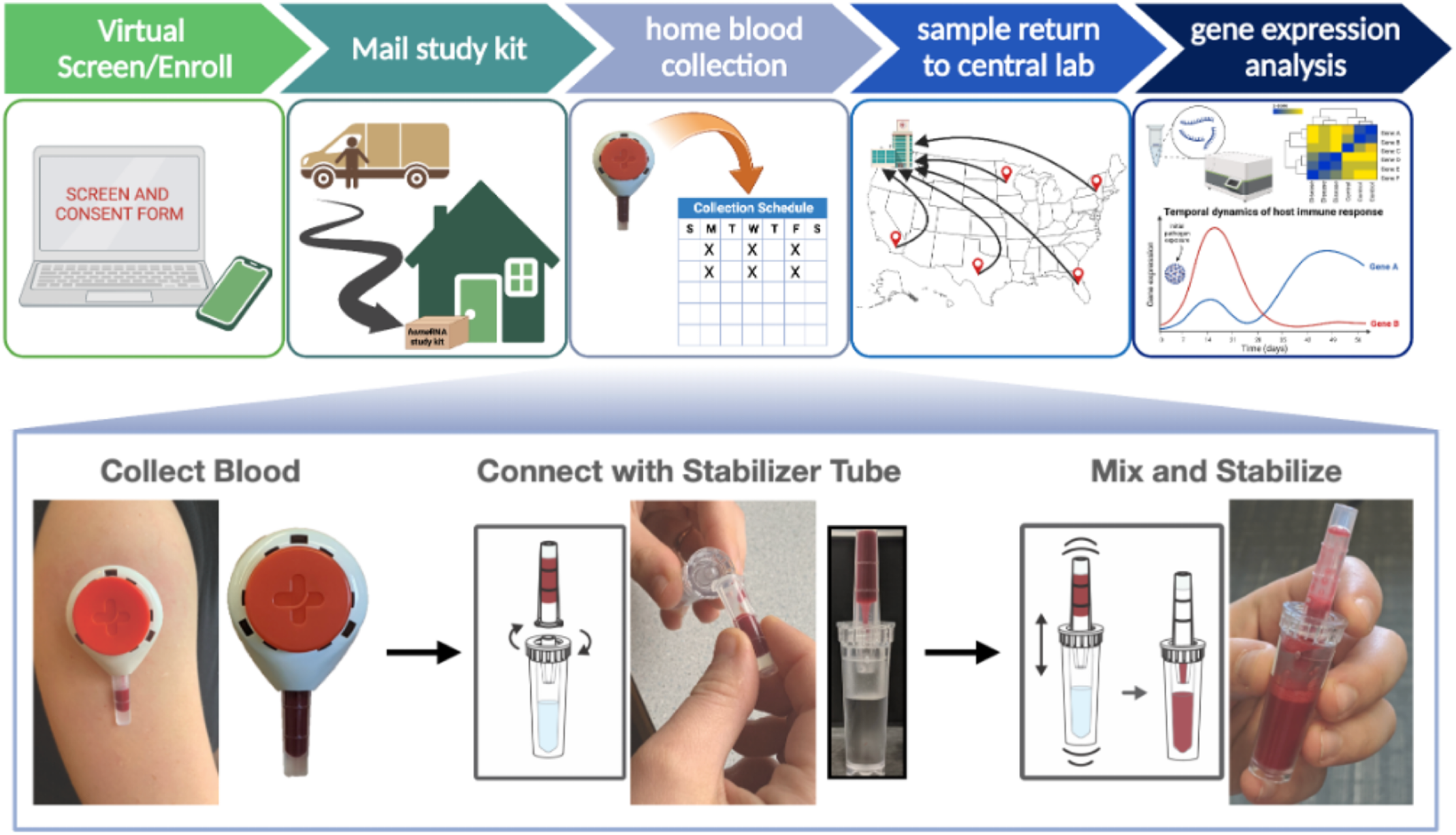
*home*RNA blood collection sampling and stabilization. Remote study design and self-blood collection from home enables decentralized sampling. *home*RNA allows the user to collect up to 0.5 mL of capillary blood (without needing to milk the collection site as is common for finger stick blood collection) using the Tasso-SST blood collection device and perform RNA stabilization themselves within minutes of collection. Part of this figure is reprinted (adapted) with permission from (*11*) Copyright 2021 American Chemical Society.

During acute viral infections, the complex interplay between various arms of the immune system gives rise to a rapidly evolving immune landscape; a delicate and time-dependent balance between pro- and anti-inflammatory molecules is crucial to control the infection (viral clearance) while maintaining inflammatory homeostasis and preventing excessive host tissue damage. Furthermore, temporal patterns of transient host biomarkers may predict clinical outcomes such as viral clearance and symptom burden. During the on-going COVID-19 pandemic, we leveraged home sampling strategies to systematically profile the temporal landscape of host and viral factors during SARS-CoV-2 infection (COVID-19 response repertoire) in non-hospitalized adults and present the results of the first application of *home*RNA in an infectious disease observational cohort.

## RESULTS

### Study design and participant characteristics

Between January – September 2021, we enrolled participants across King County, Washington to evaluate the utility of a home blood self-collection methodology (*home*RNA) for longitudinal transcriptomic studies and to test its first application in characterizing the immune response to respiratory viral infections using an outpatient SARS-CoV-2 infected cohort. First, we compared both PAXgene venipuncture and *home*RNA methodologies in healthy adult participants (*n* = 20), with RNA yield and quality as primary outcome measures (**Fig. S1A**). Next, we conducted a longitudinal observational case-control study using a SARS-CoV-2 outpatient cohort (**Fig. 2A**). Here, 39 outpatient (mild) COVID-19 participants were recruited through the Fred Hutchinson COVID-19 Clinical Research Center within 7 days of PCR positivity for SARS-CoV-2 and balanced for vaccination status (**Fig. 2A**). The median days post first symptom onset (PSO) was 9 (IQR: 8.00-10.25) for all COVID-19+ participants. Additionally, healthy uninfected participants (*n* = 5) with no history of respiratory symptoms or infection within two weeks of eligibility screen were recruited as COVID-negative controls (**Fig. 2A**). The demographics and clinical characteristics of study participants are summarized in **Table 1**.

**Table 1.** Demographics and Clinical Characteristics.

|  | Cohort B<br>(n = 35) |  |  | Cohort A<br>(n = 20) |
| --- | --- | --- | --- | --- |
|  | COVID positive (n = 30) |  | COVID<br>negative<br>(n = 5) |  |
|  | unvaccinated (n = 14) | vaccinated (n = 16) |  |  |
| Demographics |  |  |  |  |
| Sex, N (% cohort) |  |  |  |  |
| Female | 9 (25.7%) | 10 (28.6%) | 5 (14.3%) | 12 (60%) |
| Male | 5 (14.3%) | 6 (17.1%) | 0 (0%) | 8 (40%) |
| Age (years), Median (IQR) | 31 (29 - 41.5) | 36.5 (31.25 - 52.5) | 42 (39.5 - 54.5) |  |
| Ethnicity, N (% cohort) |  |  |  |  |
| Hispanic | 3 (8.6%) | 1 (2.9%) | 0 (0%) | 0 (0%) |
| Non-Hispanic | 11 (31.4%) | 15 (42.9%) | 5 (14.3%) | 20 (100%) |
| Race, N (% cohort) |  |  |  |  |
| Asian | 1 (2.9%) | 0 (0%) | 1 (2.9%) | 4 (20%) |
| Black/African American | 0 (0%) | 0 (0%) | 1 (2.9%) | 0 (0%) |
| White | 8 (22.9%) | 15 (42.9%) | 3 (8.6%) | 14 (70%) |
| Two or more races | 3 (8.6%) | 1 (2.9%) | 0 (0%) | 2 (10%) |
| Unknown | 2 (5.7%) | 0 (0%) | 0 (0%) | 0 (0%) |
| Clinical characteristics |  |  |  |  |
| Weight (lbs), Median (IQR) | 165<br>(146.8 - 193.5) | 179<br>(146.3 - 201.3) | 160<br>(142.5 - 184.5) | 155<br>(127 - 193.8) |
| Height (inches), Median (IQR) | 67.5 (66 - 69.9) | 67.5 (64.3 - 70) | 67 (59 - 71) | 66 (64 - 69) |
| Disease timeline (days), Median (IQR) [min-max] |  |  |  |  |
| post 1<sup>st</sup> symptom onset | 9 (8 - 10.25) [5 - 15] | 9 (8 - 10.75) [6 - 16] | NA | NA |
| post 1<sup>st</sup> SARS-CoV-2+ test | 7 (6 - 8) [4 - 8] | 8 (7 - 8) [6 - 9] | NA | NA |

**Figure 2.**
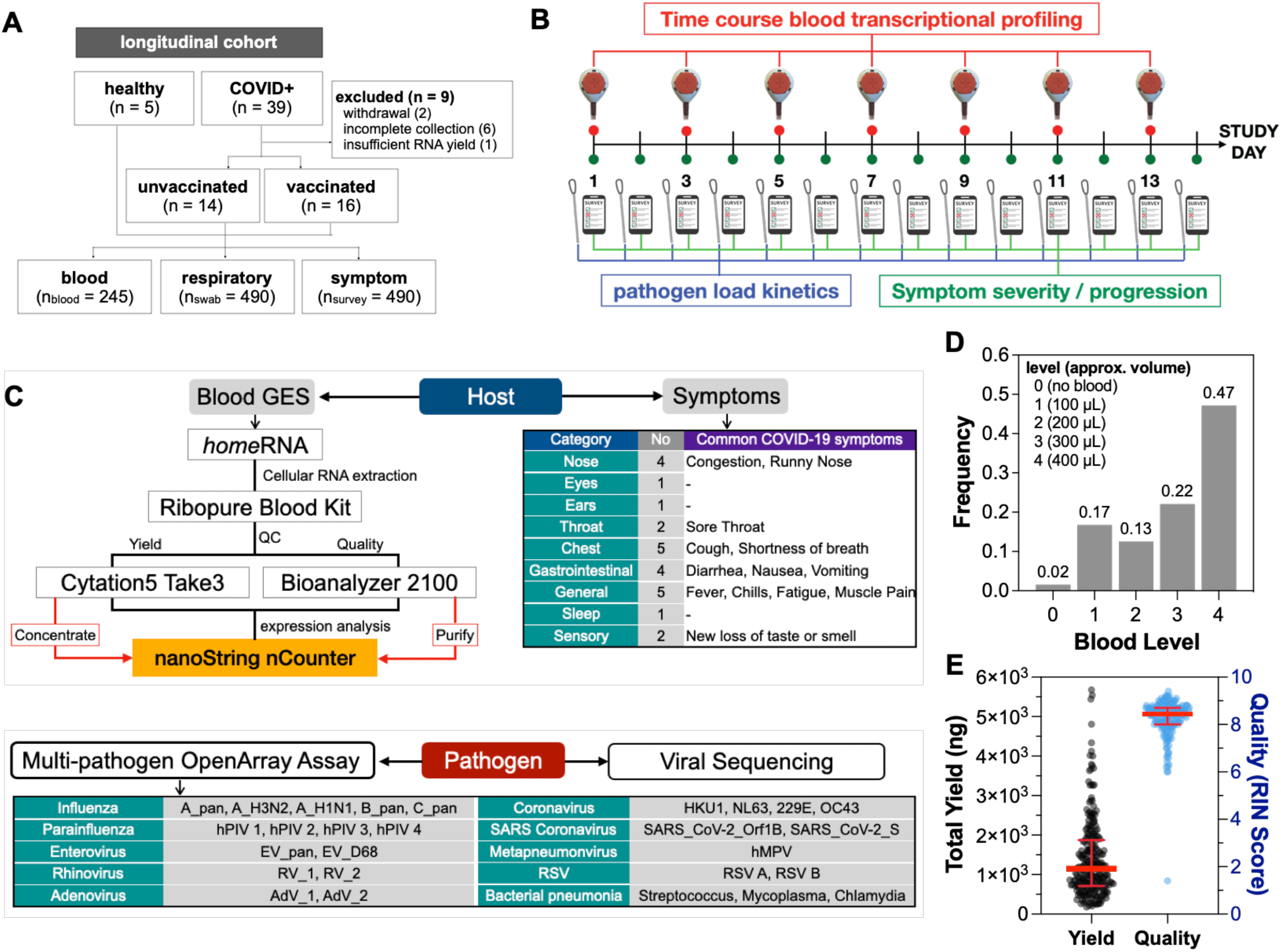
Longitudinal transcriptional profiling of the SARS-CoV-2 acute phase response. **A**) Flow chart of cohort characteristics. **B**) Study design depicting frequency of blood and nasal swab collection and symptom burden assessment. **C**) Flowchart depicting both host- and pathogen-associated outcomes measured in the study and their respective sample analysis workflows. **D**) Collected blood volume using Tasso-SST. **E**) Total RNA yield and quality of isolated RNA. Red error bars denote median with interquartile range of both RNA yield and RIN scores.

A study kit package containing seven *home*RNA blood kits and fourteen nasal swab kits was sent to all participants upon informed consent. Participants were asked to self-collect and perform stabilization of capillary blood from the upper arm using the *home*RNA blood kit every other day for a period of two weeks (**Fig. 2B**). A device use survey was distributed with each blood collection timepoint to evaluate usability and blood collection parameters. COVID-19+ participants also collected daily nasal swabs and symptom surveys to track viral load kinetics and symptom burden respectively (**Fig. 2B**). 95.5% (*n* = 44) of the participants completed the study. Longitudinal blood and nasal swab samples obtained from 35 participants were used for transcriptional profiling and pathogen analyses respectively while seven participants were excluded due to insufficient RNA yield (*n* = 1) and incomplete collections (*n* = 6) (**Fig. 2B, Table S1**). Blood transcriptional signatures were profiled for 773 genes spanning 56 immune pathways using the nCounter Host Response codeset (NanoString Inc.) (**Fig. 2C**).

### Utility of home blood collection and stabilization methodology for longitudinal transcriptomic studies

Using donor-matched samples (**Fig. S1A**), we observed comparable RNA yield and quality between PAXgene and *home*RNA sampling methodologies. As expected, PAXgene samples (2.5 mL collection volume) afforded a significantly higher total RNA yield compared to *home*RNA samples (0.1 – 0.4 mL micro-sampling volume) (**Fig. S1B and S1C, Table 2**). However, both PAXgene and *home*RNA afforded comparable yields per mL blood volume (**Fig. S1B and S1C, Table 2**). RIN scores were comparable between both sampling methodologies with *home*RNA-stabilized samples showing slightly higher RIN scores in individual samples (**Fig. S1B and S1C, Table 2**).

**Table 2.** PAXgene and *home*RNA comparison.

| RNA yield and quality measures | PAXgene (n = 20) | homeRNA (n = 19) | p-val* |
| --- | --- | --- | --- |
| Yield [total], mean (min-max) | 8.21 (4.26 - 11.8) | 0.88 (0.12 - 2.11) | <0.0001 |
| median (IQR) | 8.67 (5.06 - 10.9) | 0.82 (0.43 - 1.19) |  |
| Yield [per 1 mL blood], mean (min-max) | 3.28 (1.71 - 4.72) | 3.00 (1.05 - 5.28) | 0.478 |
| median (IQR) | 3.47 (2.02 - 4.37) | 2.94 (1.65 - 4.12) |  |
| RIN scores, mean (min-max) | 7.4 (6.6 - 8.1) | 7.8 (7.0 - 8.4) | 0.0018 |
| median (IQR) | 7.5 (7.1 - 7.7) | 7.9 (7.5 - 8.1) |  |

\*p-values were computed from an unpaired, non-parametric Mann-Whitney test.

Within our longitudinal case-control cohort (**Fig. 2A**), 36 participants completed study procedures with 98% (247/252) of the scheduled *home*RNA blood samples and 93% (482/518) of scheduled nasal swabs successfully returned to the lab (**Table S1**). Only 1% (3/252) of the *home*RNA collections failed to obtain blood while over 45% (115/252) of self-collections resulted in the maximum blood volume (**Fig. 2D**; **Table S2**). 6% (15/247) of returned blood samples did not yield sufficient RNA for transcriptomics analysis (**Table S2**). We observed a median total yield of 1.15 µg (IQR 0.71 – 1.87) and a median RIN score of 8.5 (IQR 8.0 – 8.7) (**Fig. 2E**). Of note, we successfully profiled 79% (33/42) of samples with low (∼100 µL; Level 1) reported blood collection volume (**Table S2**), demonstrating that transcriptomics applications using self-collected samples was robust even with collection volume variations.

As kit usability can affect user acceptance of this sampling methodology, we further assessed pain level, ease-of-use, blood collection time, and overall kit usage time. Over 94% of participants reported total kit usage time of under 15 minutes (**Fig. S1D**) and completed blood collection within 6 minutes (**Fig. S1E**). Similarly, over 90% of participants reported slight to no pain and ease of use for both the Tasso-SST blood collection and RNA stabilizer tubes (**Fig. S1F**).

### Viral load kinetics and symptom burden in an outpatient SARS-CoV-2 cohort

To track SARS-CoV-2 viral load (VL) kinetics and detect co-infections with other respiratory pathogens, nasal swabs from COVID-19+ participants were subjected to a multi-pathogen RT-qPCR assay on the OpenArray platform (Thermo Fisher) (**Fig. 2C**). Swabs with time-matched blood samples were serially tested until we observed clearance of all detected respiratory pathogens in two consecutive samples (**Fig. 3A**). As shown in **Fig. 3B**, single cases of co-infection with adenovirus and *Streptococcus pneumoniae* were detected while 20% (6/30) of COVID-19+ participants in our cohort were co-infected with human rhinovirus (**Fig. 3B**). SARS-CoV-2 sequencing showed infections from five variants (Alpha, Epsilon, and Delta) including ancestral lineages 20A and 20B (**Fig. 3B**). Due to the enrollment timeline, we observed a higher representation of both Alpha and Epsilon variants in unvaccinated individuals compared to vaccinated individuals, who were predominantly (56%) infected with the Delta variant (**Fig. 3B**; **Table S3**). We were not able to obtain variant information on four participants (3 vaccinated and 1 unvaccinated) due to low or no viral load (**Fig. 3B**). When viral loads were aligned to days PSO, we observed that unvaccinated individuals presented with higher viral loads and delayed viral clearance compared to vaccinated individuals; this difference did not reach statistical significance (**Fig. 3C and 3D**). Unvaccinated participants showed delayed viral clearance (16.5 PSO; IQR 12.75-19.5) compared to breakthrough infections (13.5 PSO; IQR 10.25-15.75), pointing towards a longer period of contagiousness (**Fig. 3D**). From participant symptom surveys, more than 30% of COVID-19+ participants experienced altered sensory (smell and taste), general fatigue, and respiratory congestion (**Fig. 4A**), with altered smell and taste having the highest symptom severity scores (**Fig. 4B**). Although we did not observe significant differences in symptom severity between unvaccinated and vaccinated participants, we noted an overall milder presentation of respiratory symptoms in vaccinated individuals (**Fig. 4C**). Symptom severity for individual participants is depicted in **Fig. S2**. Spearman correlation analysis between viral load and symptom burden showed a strong positive correlation between viral load, altered smell and taste (S1/S2), and disrupted sleep (H3). (**Fig. 4C and 4D**).

**Figure 3.**
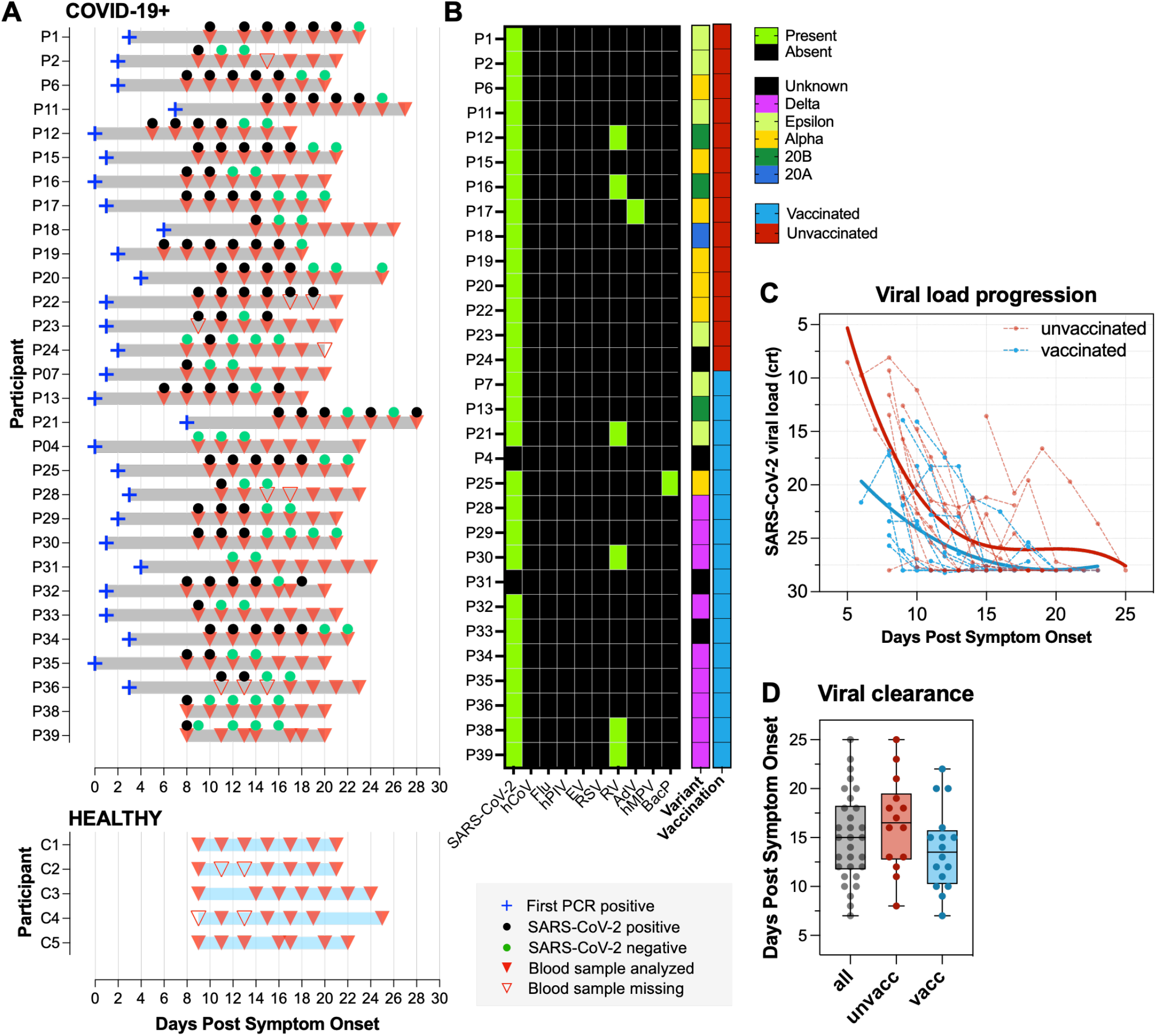
SARS-CoV-2 viral load kinetics in unvaccinated and vaccinated COVID-19+ participants. **A**) Disease timeline and participant blood and nasal swab samples aligned to days PSO (Day 0) in both COVID-19+ participants and healthy controls. Blue cross denotes first PCR positive day; black and green circles denote SARS-CoV-2 positive and negative nasal swab samples respectively; solid red triangles denote blood samples used in gene expression analysis while transparent red triangles denote missing blood samples. **B**) Heatmap depicting presence of SARS-CoV-2 and other respiratory pathogens in participants’ nasal swab samples. Right annotation columns depict sequenced SARS-CoV-2 variants and participants’ vaccination status. **C**) SARS-CoV-2 viral load progression. Dotted lines represent individual viral load trajectory. Solid lines represent the loess smooth function of viral load within each vaccination subgroup. **D**) Box and whisker plot depicts viral clearance in unvaccinated versus vaccinated COVID-19+ participants.

**Figure 4.**
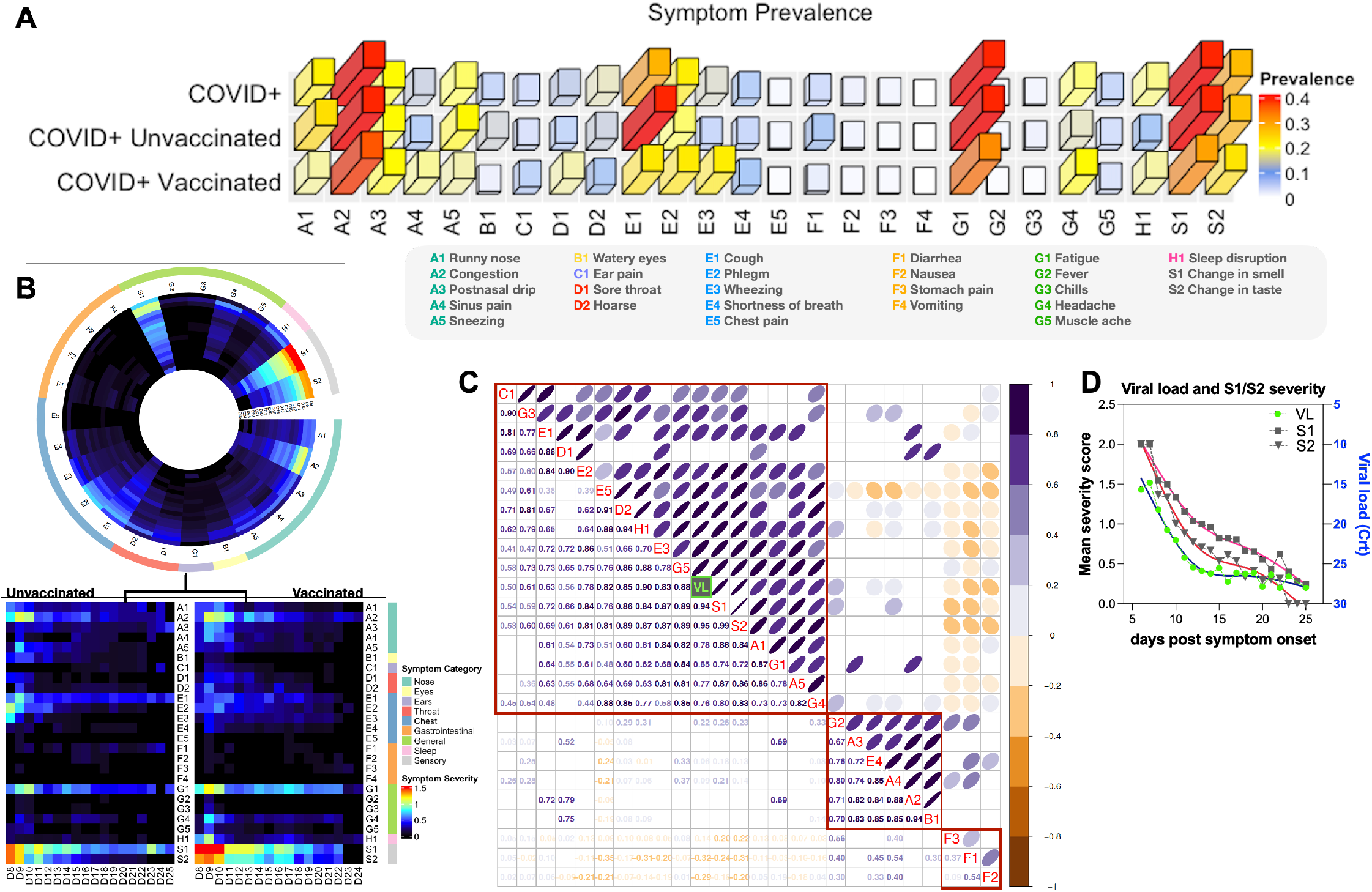
Correlation between viral load and symptom severity in COVID-19+ participants. **A**) 3D bar plot depicts symptom prevalence (relative abundance) of 26 surveyed symptoms across 9 broad categories (A-S). **B)** Circos heatmap depicts mean symptom severity score for each symptom type from days 8-25 PSO (rows) in COVID-19+ participants. Red denotes high symptom severity score while black denotes no experienced symptoms. Outer circle depicts broad categories of individual symptoms. Linear heatmap stratifies symptom severity scores by participants’ vaccination status. **C**) Spearman correlation analysis of SARS-CoV-2 viral load and symptom severity. Mean viral load for each day PSO and mean symptom severity score for each symptom type in COVID-19+ participants were analyzed. Purple and yellow denotes positive and negative correlation respectively. Width of the circle in the upper right matrix denotes the strength of the association with strongest association depicted by a thin line. Bottom left matrix depicts the Spearman correlation coefficient. Blank cells denote no statistical significance for that particular association. **D**) Mean viral load and S1/S2 severity scores across all COVID-19+ participants aligned to days PSO.

### At-home self-collection and stabilization of capillary blood captures host transcriptional responses to SARS-CoV-2 infection in an outpatient cohort

We analyzed a total of 232 longitudinal capillary blood RNA samples from 30 COVID-19+ and 5 healthy participants (**Fig. 2A**) and 6 single timepoint samples obtained from healthy participants (**Fig. S1A**). Using nCounter direct digital counting of native mRNA, we profiled expression of 773 genes spanning 56 biological pathways involved in the host immune response to infectious diseases. All 238 assayed participant samples passed the manufacturer’s recommended QC parameters (**Fig. S3**) and were normalized for i) assay variations using positive control spike-ins, ii) codeset versions using manufacturer-provided panel standards, and iii) sample input using nine reference genes. We fitted generalized additive mixed models (GAMMs) to the normalized counts to describe the kinetics of gene expression and examine their potential association with disease and/or vaccination status (**Table S4**; **Fig. 5A**) (*12*). We fitted three models: **Model 1** assessed differentially expressed genes (DEGs) between COVID-19+ and healthy participants [*covid19:healthy*]; **Model 2** assessed DEGs between vaccinated and unvaccinated COVID-19+ participants [*covid19 vacc:unvacc*] using only samples derived from COVID-19+ participants; **Model 3** contrasted COVID-19+ vaccination subgroups to the healthy reference group [*covid19 vacc:healthy* and *covid19 unvacc:healthy*]. All three models adjusted for age, sex, and host response codeset version as covariates (**Table S4**). In addition to pairwise contrasts, we fitted smooth functions in all models using days PSO as a continuous variable [*s(days)*] to identify temporally dynamic genes in four major participant groups: healthy [*s(days):healthy*], COVID-19+ unvaccinated [*s(days):unvacc*], COVID-19+ partially vaccinated [*s(days):vacc(partial)*], and COVID-19+ vaccinated [*s(days):vacc(full)*] (**Fig. 5A**). As all healthy participants within our cohort are vaccinated, we introduced a fourth category level for vaccination status (Healthy) in **Model 3** to allow us to distinguish responses by disease status (**Table S4**). **Table S4** summarizes, for each fitted GAMM, the confounder adjustments, categorical variables used for vaccination status, samples fitted within each model, and the pairwise contrasts and smooth functions obtained from each model. An adjusted p-value < 0.1 using the Benjamini-Hochberg procedure is used to call significant genes.

**Figure 5.**
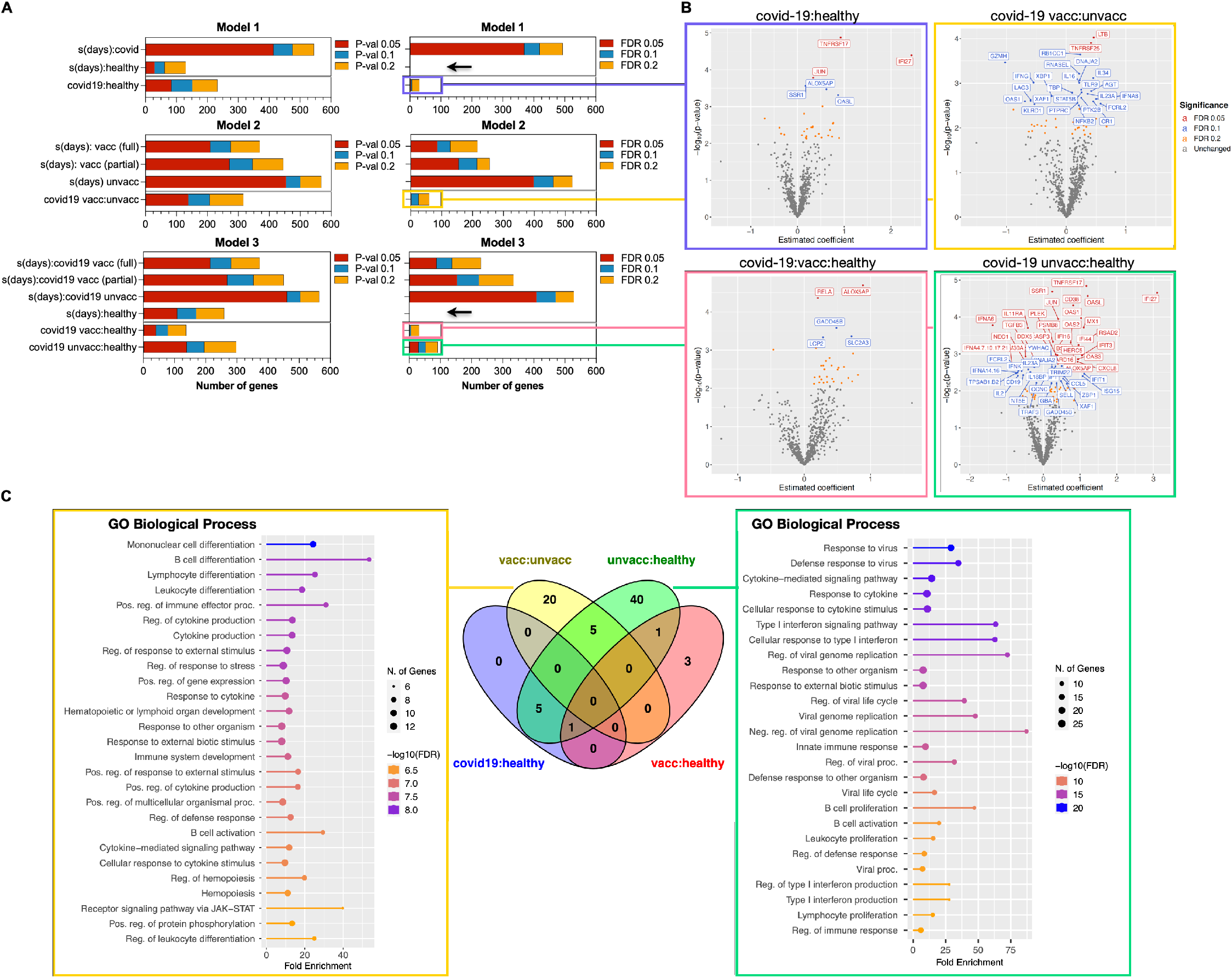
Differentially expressed genes (DEGs) in COVID-19+ vaccinated and unvaccinated participants. **A**) Stacked bar plots depict number of DEGs from all three fitted GAMMs. Left and right plots depict the number of genes with raw and adjusted (FDR) p-values < 0.05, 0.1, and 0.2 respectively. Gray lines within each plot separate results of smooth function of days PSO [*s(days):group of interest*] and pairwise contrasts [*test group:reference group*]. Black arrow highlights lack of significant dynamic genes identified in healthy uninfected participants. **B**) Volcano plots depict significant DEGs from pairwise contrast groups shown in A). Genes are colored based on FDR cutoffs. Significant genes (FDR < 0.1) are labeled. x-axis depicts the estimated coefficient fitted from its respective GAMM while y-axis depicts the -log10(raw p-value). **C**) Venn diagram depicts gene overlap of significant DEGs in each pairwise contrast groups. Lollipop plots depict GO functional enrichment of genes unique to the [*vacc:unvacc*] and [*unvacc:healthy*] contrast groups.

Pairwise contrast analyses between disease and vaccination subgroups showed a low number of significantly DEGs in both the [*covid19:healthy*] (6 total DEGs) and [*covid19 vacc:healthy*] (5 total DEGs) contrast groups, followed by [*covid19 vacc:unvacc*] (25 total DEGs) and [*covid19 unvacc:healthy*] (52 total DEGs) when false discovery rate (FDR) is controlled at 10% (**Fig. 5A** and **5B**). Functional enrichment of the 40 DEGs unique to [*covid19 unvacc:healthy*] using the Gene Ontology (GO) Biological Process database showed enrichment of pathways involved in antiviral responses, cytokine signaling and responses, Type I interferon signaling and responses, and B cell activation and proliferation (**Fig. 5C**). On the contrary, 20 of the unique DEGs in [*covid-19 vacc:unvacc*] showed functional enrichment in pathways associated with various immune cell differentiation (e.g., mononuclear, B-cell, lymphocyte, leukocyte, etc.) and regulation of cytokine production and stress response (**Fig. 5C**). Taken together, our results demonstrate that at-home self-collection and stabilization of capillary blood using *home*RNA is capable of capturing key acute phase antiviral transcriptional responses in mild SARS-CoV-2 cases (outpatient setting) and suggest a distinct acute phase response between unvaccinated and breakthrough infections. Notably, we did not observe any significant dynamic genes in the healthy controls in both Models 1 and 3 (**Fig. 5A**).

### Mining dynamic genes through longitudinal transcriptional profiling provides insights into the COVID-19 acute phase response repertoire

Frequent and systematic sampling intervals over a 2-week period enabled us to profile the host immune kinetics at high-resolution during acute SARS-CoV-2 infection. By incorporating smooth functions [*s(days)*] into our models, genes with significant temporal dynamicity (response repertoire) were identified within each disease status and vaccination subgroup (**Table S4**; **Fig. 5A**). Using **Model 1**, we aimed to identify dynamic genes in COVID-19+ participants [*s(days):covid19*] and contrasted this list against the healthy reference group [*s(days):healthy*]. We identified a total of 418 dynamic genes (adjusted p-value < 0.1) within the COVID-19+ response repertoire (**Fig. 5A**). On the contrary, we observed no dynamic genes across the same temporal window within the healthy reference group (**Fig. 5A**). As vaccination can reprogram the host response to infection, we further stratified the smooth function analyses of COVID-19+ response into three vaccination subgroups using **Model 3 (Table S4**; **Fig. 5A**). We observed that unvaccinated COVID-19+ individuals [*s(days):unvacc*] presented the largest response repertoire consisting of 470 dynamic genes compared to vaccinated individuals [*s(days):vacc*] with a total response repertoire of 137 dynamic genes (**Fig. 5A**). Corroborating **Model 1**, no dynamic genes were observed in the healthy reference group in **Model 3** (**Fig. 5A**). Comparison of response repertoire memberships between COVID-19+ and its respective vaccination subgroups revealed that a significant proportion of the COVID-19+ response within our cohort is driven by unvaccinated participants (**Fig. 6A**). A majority of the vaccinated subgroup repertoire consisted of genes with shared membership across all three comparison groups (**Fig. 6A**). Interestingly, smooth function analyses identified a subset of 96 response genes unique to the unvaccinated response repertoire functionally enriched in leukocyte and myeloid activation genes (**Fig. 6B**). When hierarchical clustering was performed on the top-ranked (by adjusted p-value) dynamic genes (*n* = 100) identified from the COVID-19+ unvaccinated response repertoire using samples obtained from the first sampling timepoint, a distinct transcriptional signature that consists of i) higher expression levels of Type I interferon (**Fig. 6C**; left panels C, J, and K), Type II interferon, and RNA sensing genes ii) lower expression of myeloid activation and TLR/TCR/BCR signaling genes (**Fig. 6C**; left panels A, E-G), and iii) enrichment of cytotoxic cell markers (**Fig. 6C**; left panel L) clustered COVID-19+ unvaccinated subgroups from vaccinated counterparts, whose transcriptional signatures more closely paralleled that of the healthy reference group (**Fig. 6C**).

**Figure 6.**
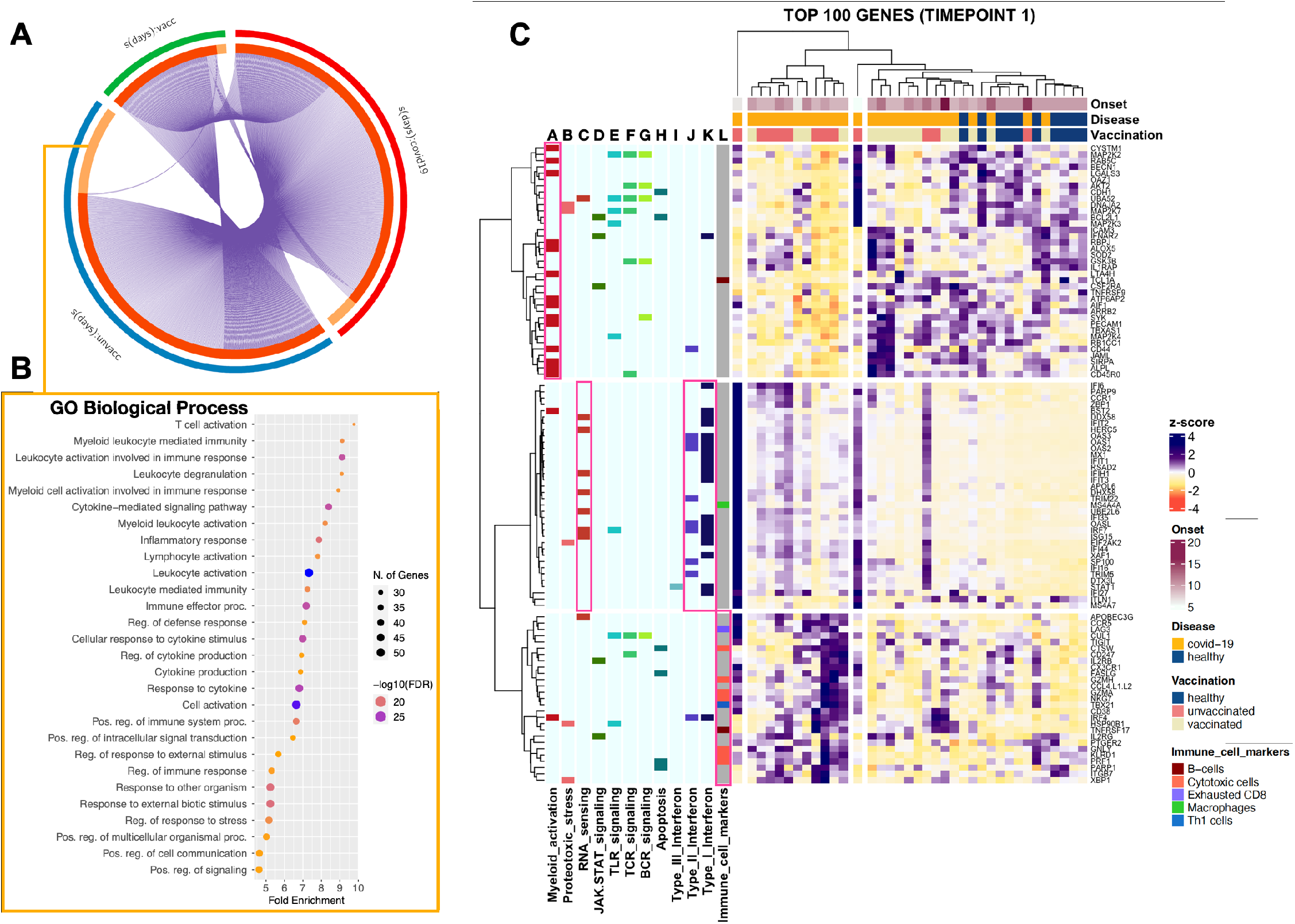
The COVID-19 response repertoire of mild-to-moderate outpatient infections. **A**) Circos plot showing overlap of significant genes identified from GAMM smooth function analysis between unvaccinated and breakthrough infections. Purple lines link identical genes between each group. The inner circle represents gene lists, where hits are arranged along the arc (dark orange denotes genes that hit multiple lists; light orange denotes genes unique to a particular list). **B**) Dot plot showing functional enrichment of genes unique to the COVID-19 unvaccinated response. The size of the dot represents number of enriched genes. **C)** Heatmap depicting hierarchical clustering (Euclidean Ward D2) of top 100 ranked dynamic genes identified from the COVID-19 unvaccinated response repertoire, where rows represent genes and columns represent the first timepoint sample from each participant. Top horizontal bars represent clinical characteristics of the corresponding samples. Left vertical bars represent select Host Response pathway annotation (light blue columns) and annotated immune cell markers (gray column). Various solid colors depict positive membership of a particular gene within that pathway.

### Time-course geneset analysis depicts a robust interferon response in COVID-19+ unvaccinated subgroup

To further assess dynamics of co-expressed genes networks, we performed a time-course geneset analysis (TcGSA) comparing COVID-19+ samples to the healthy reference group using a highly curated blood transcriptional response geneset module (Chaussabel BloodGen3 Modules) (*13-15*) (**Fig. 7A**). Due to the variations in disease onset timeline across all participants, dynamic genesets were assessed across seven total sampling timepoints and an adjusted p-value < 0.1 using the Benjamini-Hochberg procedure was used to call significant genesets. To evaluate if temporal dynamics between the two vaccination subgroups was comparable when using sampling timepoints instead of days PSO, we plotted each timepoint sample between the two major vaccination subgroups with days post symptom onset on the x-axis (**Fig. 7B**). As shown in this figure, we observed no significant difference between the median days PSO between the two vaccination subgroups across all timepoints. We identified 12 dynamic genesets when COVID-19+ participants were compared to reference healthy groups (**Fig. 7A**). Supporting the above results, TcGSA estimations showed a robust interferon response coupled with cytotoxic lymphocyte that waned over time. On the contrary, we observed increased expression of inflammation and B-cell related genesets at later sampling timepoints (**Fig. 7A**). **Figure 7C** depicts the temporal dynamics of three top-ranked dynamic genesets (M8.3, M9.1, and M15.127) along with their gene membership composition. Both interferon modules M8.3 and M15.127 showed tight homogenous co-expression between all module membership genes. The cytotoxic lymphocyte module (M9.1) showed higher heterogenicity in gene expression within the module. In the healthy reference group, both interferon modules showed stable kinetics while a very robust response was observed in early timepoints in the COVID-19 unvaccinated subgroup. Vaccinated COVID-19+ participants did not mount a robust interferon response within this captured temporal disease window.

**Figure 7.**
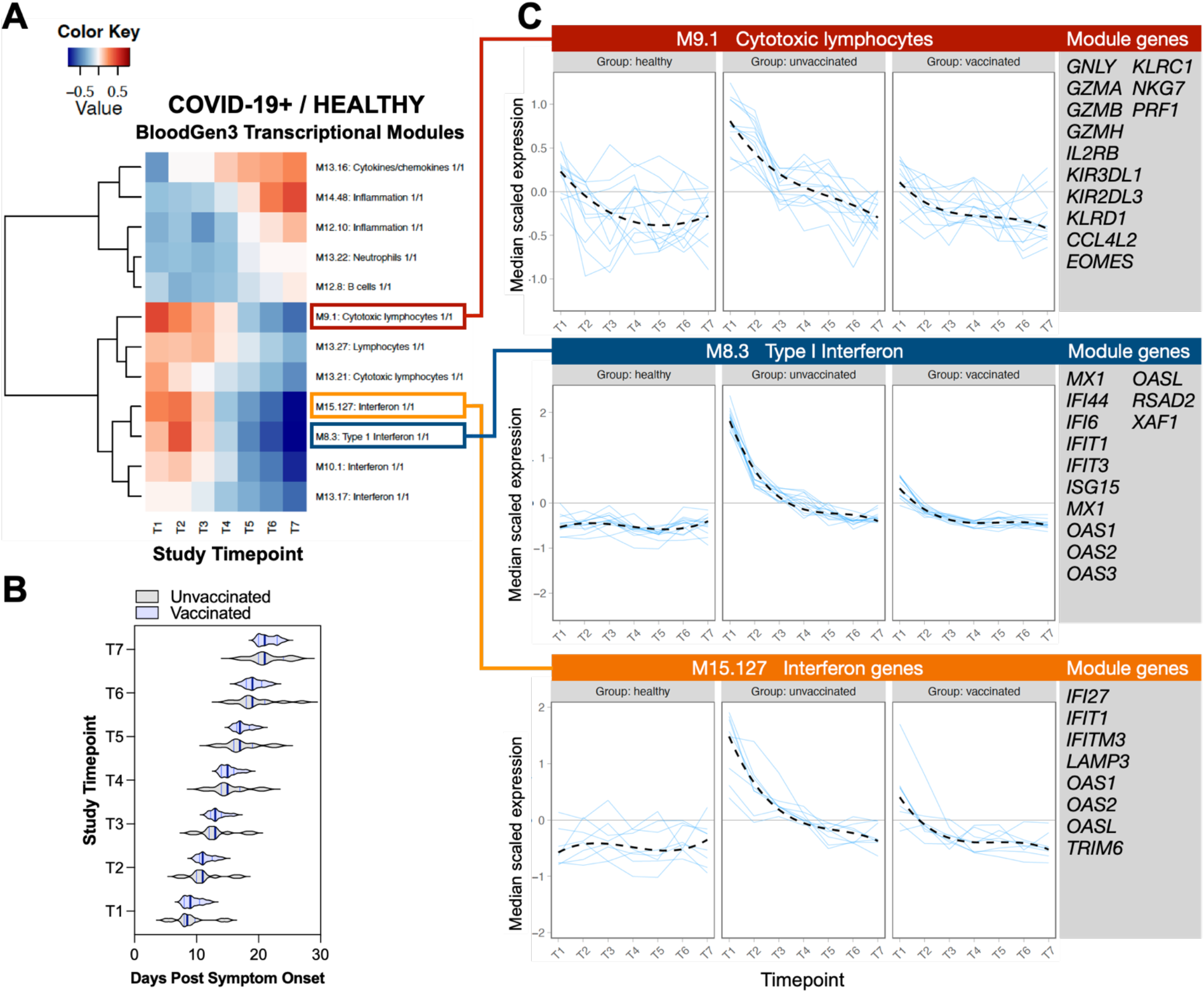
Time-course geneset analyses (TcGSA) reveals higher interferon response intensity in unvaccinated individuals. **A**) Heatmap depicting expression kinetics of significant genesets identified from TcGSA. Chaussabel BloodGen3 modules were used to query all genes within our dataset. Columns represent sampling timepoints 1-7 while rows represent significant geneset modules. **B**) Violin plot compares median days PSO between unvaccinated and vaccinated subgroup in each sampling timepoint. **C**) Spaghetti plots depict expression kinetics (median scaled gene expression) of individual genes (blue line) within each module in healthy, unvaccinated, and breakthrough COVID-19 infections. Gene memberships are listed to the right of each module.

### The interferon-response gene, *IFI27*, tracks closely with viral kinetics

To visualize kinetics of dynamic genes identified from either GAMMs or TcGSA analyses above, we plotted expression of select genes for each individual participant (**Fig. 8A**). As shown in **Fig. 8A**, the smooth function fit for all positive and negative controls in addition to four representative reference genes did not show temporal dynamicity for both COVID-19+ and healthy groups (**Fig. 8A**). On the contrary, we observed several temporal trends for dynamic genes identified from previous GAMMs and TcGSA analyses (**Fig. 8A**). Genes within the cytotoxic lymphocyte module M9.1 (**Fig. 7C**) such as *PRF1* and *KLRD1* showed slight increase in expression in COVID-19+ compared to healthy controls that gradually waned over the course of the observed temporal window. On the contrary, genes within the interferon response modules (M8.3 and M15.127) such as *ISG15, XAF1, IFI27*, and *IFIT1* showed significantly elevated expression in COVID-19+ compared to healthy controls early in the observed window. Unlike the gradual waning of expression over time, expression of these genes decreased dramatically and returned to presumed baseline levels around 15 days PSO (**Fig. 8A**). We observed that *IFI27* displayed the largest temporal change in COVID-19+ participants. We next assessed if expression of *IFI27*, an interferon response gene displaying the highest increase in expression, correlated with viral clearance and symptom severity on a participant-specific level. To achieve this, we plotted i) symptom severity and number, ii) SARS-CoV-2 viral load, and ii) *IFI27* gene expression for each individual participant aligned to days PSO (**Fig. 8B** and **Fig. S4**). We observed that the temporal kinetics of *IFI27* tracks closely to that of SARS-CoV-2 viral load regardless of vaccination status of the COVID-19+ participants (**Fig. 8B**).

**Figure 8.**
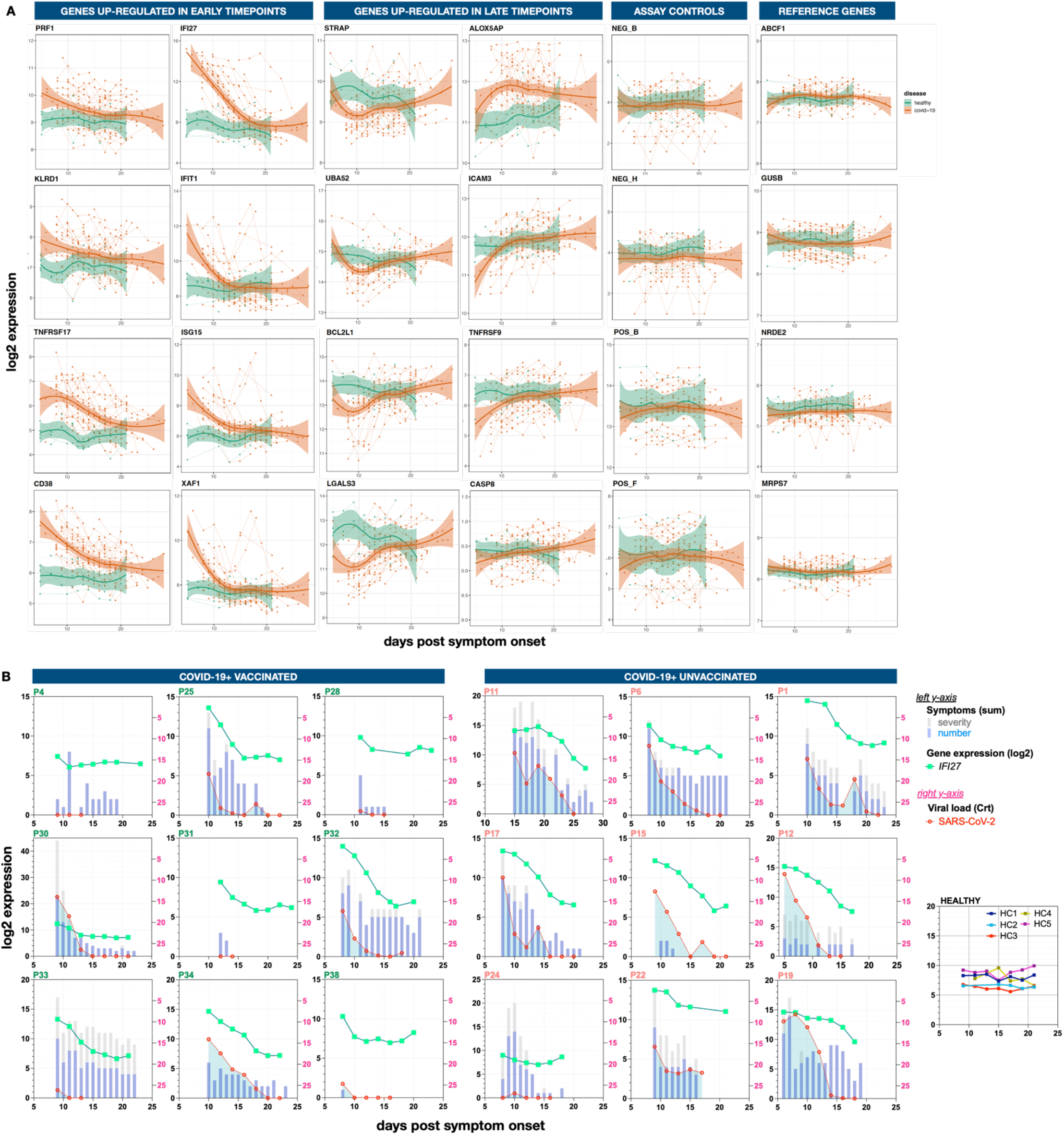
*IFI27* expression tracks with SARS-CoV-2 viral load. **A**) Temporal trends of select response genes upregulated in early and late timepoints in COVID-19+ participants. Dotted lines connecting each analyzed sample (solid circles) represent expression kinetics in individual participants. Solid lines represent estimated loess smooth function in COVID-19+ participants (red) and healthy infected controls (green). Shaded area depicts confidence intervals. **B**) Correlation between *IFI27* gene expression, viral load, and symptoms in individual participants. Total symptom number (blue columns), total symptom severity (gray columns), and *IFI27* gene expression kinetics are plotted on the left *y-axis*. SARS-CoV-2 viral load is plotted on the right *y-axis. IFI27* expression kinetics of all healthy uninfected controls are displayed on the far-right plot. Scale for left y-axis adjusted to symptom number and severity.

In addition to interferon and innate antiviral genes, we observed a group of co-expressed genes that show increased expression throughout the observation window. **Fig. 8A** depicts four select dynamic genes (*ALOX5AP, ICAM3, TNFRSF9*, and *CASP3*) identified from both GAMMs and TcGSA analyses. Corroborating results from previous smooth function analyses (**Fig. 5A**), healthy uninfected controls showed steady expression levels of these genes over time. Intriguingly, by performing a high frequency systematic sampling of SARS-CoV-2 acute phase response, we observed a group of genes (select four *STRAP, UBA52, BCL2L1, LGALS3* shown in **Fig. 8A**) displaying a transient dip in expression early in the observation window, characterized by rapid suppression of gene expression followed by a rapid recovery to observed baseline level (based on our sampling window) as early as one week post onset of the suppression (e.g. *BCL2L1* and *LGALS3*). Upon returning to observed baseline, expression of these genes continued to increase throughout the observation window (**Fig. 8A**). This transient and rapid pattern in dynamicity of the immune response during acute infection would be missed in single timepoint sampling or even longitudinal designs that are limited to weekly sampling due to barriers to sample collection. *LGALS3*, encoding for the β-galactoside-binding protein (Galectin-3), has been shown to mediate viral attachment and entry in herpes simplex virus infection (*16*) and enhance viral infection (e.g. influenza) and airway inflammation (*17*). Playing a pro-inflammatory role in driving neutrophil infiltration and airway inflammation, *LGALS3* correlates with severe outcome in COVID-19 infection (*18*). The observed transient systemic suppression of *LGALS3* may provide protective homeostasis against damaging airway inflammation.

## DISCUSSION

Whole blood transcriptional profiling is a powerful tool for the evaluation of immune responses to infectious diseases; however, blood collection via traditional phlebotomy remains a barrier to precise characterization of the immune response in highly dynamic infections such as those caused by respiratory viruses (*8, 19*). Here we present our study using an at-home self-collection methodology, *home*RNA, to capture temporal gene expression profiles during the acute phase of mild-to-moderate SARS-CoV-2 infection. Our data demonstrate that home-based blood self-collection in the setting of acute illness is feasible, with high participant usability scores, and yields sufficient RNA quantity and quality to perform transcriptomic profiling. We also showed that longitudinal gene expression profiles vary significantly between COVID-19 and healthy participants, especially when comparing unvaccinated infected individuals to healthy controls. Furthermore, longitudinal blood sampling coupled with symptom surveys and viral load data revealed temporal relationships between host immune responses, symptoms, and viral load kinetics, especially with respect to interferon genes such as *IFI27*.

Home-based RNA preservation platforms like *home*RNA are particularly beneficial in the setting of highly communicable diseases such as SARS-CoV-2, where in-person phlebotomy visits are not desirable given the need for strict isolation measures and high exposure risk to healthcare workers; in-person phlebotomy, especially in the context of longitudinal sampling designs, is also highly inconvenient for infected individuals. Decentralized blood self-collection also enables on-demand and systematic distribution, collection, stabilization, and return of liquid blood specimen and is scalable to geographically disparate multi-site longitudinal studies. The temporal flexibility in both the timing and frequency of sampling also allows capture of transient but highly dynamic transcriptional signatures at specific treatment or disease stages. Previous studies of home-based blood collection for RNA stabilization employ a fingerprick followed by capillary tube transfer into microcentrifuge tubes containing RNA preservation medium (*20, 21*). The *home*RNA kit allows for collection of larger volumes of blood compared to the fingerprick approach (400ul with *home*RNA versus 50ul with fingerstick), thus yielding more RNA for omics-based applications (**Fig. 1**).

A critical metric for home-based blood self-collection for transcriptomics is reproducibility and stability of expression signatures over time despite variations in specimen storage and transit durations. We demonstrated our ability to achieve this with *home*RNA in two ways. First, we showed that individual housekeeping genes are expressed at stable levels over time in both healthy and COVID+ subjects (**Fig 8A**). Secondly, healthy uninfected controls showed remarkably stable expression trends over time, with no differentially expressed genes over time (dynamic genes) in co-variate-adjusted smooth function models (**Fig 5A**). This is further demonstrated at the gene module and individual gene level, where we observed stable expression trends in healthy individuals of several genes and co-regulated pathways associated with interferon responses (**Fig 7C** and **8A**). These findings in aggregate suggest that our platform is reliable and reproducible for longitudinal sampling.

To the best of our knowledge, this is the first study to evaluate longitudinal gene expression signatures correlated to time-matched viral load kinetics and symptom assessment in SARS-CoV-2 infected individuals with mild-to-moderate disease using a fully remote study design. A significant proportion of SARS-CoV-2 infections are mild cases that do not require hospitalization. Regardless, mild cases of COVID-19 can present a wide range of symptom severity and individuals with mild infections can still develop post-acute sequelae of COVID-19 (PASC) or long COVID (*22*). To date, transcriptional profiling of the immune response, especially in longitudinal studies, is heavily focused on hospital-acquired samples and controlled human challenge studies (*23-25*), oftentimes from patients with moderate-to-severe symptoms or requiring oxygen support (*26*). When mild outpatient cases are evaluated, single time-point samples are collected through initial clinic visit or resource-intensive mobile phlebotomy (*27*) and longitudinal studies are oftentimes limited to infrequent sampling timepoints (*28, 29*). One study in outpatients with asymptomatic to moderate SARS-CoV-2 collected blood for RNA-seq at days 0 and 5 post-enrollment, which was at a median of 5 days post symptom onset (*28*). Although extremely informative, transcriptional profiles of severe infection cases may not be representative of the host immune response in mild outpatient cases and thus to date, the precise characterization of the kinetics underlying the immune response to mild infections is scarce and broad transcriptional response in mild outpatient cases is not well understood.

Using *home*RNA, we can decentralize specimen self-collection and profile the host transcriptional response in an outpatient setting and can collect blood at much higher temporal frequency. Due to the variable nature of remote self-sampling pipeline, we sought to employ gene expression analyses methods that mitigate amplification of noise potentially introduced into the self-sampling technique. The molecular barcoding chemistry and single molecule digital imaging of the nCounter (nanoString) hybridization-based gene expression analysis allows for direct and amplification-free targeting of native RNA transcripts. The lack of reverse-transcription, amplification, and enzymatic steps coupled with direct probe-based hybridization of native RNA targets reduce amplification of error introduced at each processing step, allowing for highly sensitive and reproducible gene expression counts within our longitudinal dataset.

Our data demonstrate that COVID-19+ participants have a robust interferon response in the early stages of infection, most apparent among unvaccinated individuals. This finding is consistent with other studies examining gene expression signatures in the peripheral blood, where early timepoints are associated with high levels of interferon response genes that decline in samples taken mostly from unique individuals at later timepoints (*19*). Importantly, elevated *IFI27* expression has been consistently demonstrated in both early/mild and severe SARS-CoV-2 infections and can discriminate between infected and uninfected individuals (*26, 30*). *IFI27* (interferon alpha-inducible protein 27 or *ISG12a*) belongs to a family of interferon-stimulated genes whose expression has been associated with other viral infections (*31, 32*) including progression of HIV-1 (*33*) and as an early biomarker of influenza (*34*). Thus, our findings of differential expression of *IFI27* in COVID-19 infected individuals have biological relevance and is consistent with prior studies. Furthermore, our study design allowed for correlation between *IFI27* expression, viral load, and symptom severity and duration. We previously demonstrated that self-sampling with foam nasal swabs provides a quantitative viral output that correlates with nasal cytokine levels in non-SARS-CoV-2 respiratory viruses (*35*). Here we demonstrated that similar correlations between symptoms, viral load, and *IFI27* expression in the peripheral capillary blood are also present during acute SARS-CoV-2 infection.

As the immune response to viral infections can vary significantly between individuals, we assessed if at-home longitudinal self-sampling can detect distinct molecular signatures between disease sub-groups. Given that breakthrough cases (vaccinated) of COVID-19 generally present milder symptoms, we evaluated the transcriptional response in COVID-19 participants stratified by vaccination status. In a recently published study, transcriptional profiling of nasal swab samples comparing unvaccinated versus vaccinated breakthrough cases of COVID-19 demonstrated decreased innate and inflammatory response and increased adaptive host response in the breakthrough cases (*36*). Despite the lower number of study participants in our pilot dataset, we observed a parallel host transcriptional response, characterized by decreased interferon response in the periphery (**Fig. 6C** and **7C**), demonstrating that at-home longitudinal capillary sampling can preserve distinct transcriptional signatures between disease subtypes.

There are several limitations of our study. First, participants were enrolled up to 7 days following a positive SARS-CoV-2 infection, and first sampling timepoint obtained up to 16 days post symptom onset; therefore, early responses to infection are not captured in our data. Careful interpretation of temporal dynamics must be employed when interpreting the magnitude of our observed responses as response present outside of the captured temporal window may not be reflected in the dataset. For example, within our dataset, we observe a significantly lower interferon response from vaccinated COVID-19+ individuals. However, during the limited 2-week snapshot window we acquired, we may have missed an earlier but more robust interferon response mounted in vaccinated COVID-19+ individuals. Second, as home (decentralized) self-collection methodologies can be highly susceptible to technical variabilities from sample collection, stabilization, storage, and transit period, we expected some level of technical noise in the temporal dynamics. However, we observed surprisingly stable kinetics of the 773 host response genes analyzed within our healthy reference group unlike the highly dynamic transcriptional landscape of the COVID-19 acute phase samples, suggesting highly robust collection and stabilization using our sampling methodology. Third, as with other bulk transcriptomics-based analysis, our gene expression analysis method is not able to distinguish between active transcriptional remodeling versus changes in blood immune cell type composition, resulting in differential level of cell-type specific transcripts. *In silico* cell-type deconvolution algorithms such as CIBERSORTx can be applied to this dataset to further delineate transcriptional versus cell type abundance changes (*19, 37, 38*). Future development of home-based sampling methodology that is compatible with single-cell analyses platforms is of utmost interest to our group. Finally, due to our limited sample size and enrollment period, we were not able to characterize SARS-CoV-2 variant specific gene expression signatures. Clinical data suggest disease severity varies significantly by variant, especially when comparing ancestral strains and early variants to currently circulating Omicron lineage variants, and limited data suggest host immune responses can vary between strains (*39, 40*). Future studies with adequate sample sizes are needed to account for variant specific host gene expression variability.

In sum, we present the first application of a novel home-based blood collection system for profiling the host gene expression kinetics during acute phase SARS-CoV-2 that produced robust longitudinal results and demonstrate its capability of profiling transient host response mechanisms during dynamic disease stages. We believe this tool can be applied broadly to a wide repertoire of disease states, especially those with highly dynamic host gene expression profiles, such as in infectious diseases where early host responses may be predictive of disease severity.

## MATERIALS AND METHODS

### Study Design and Participant Characteristics

The objective of this study was to test the application of a home-use blood sampling and RNA stabilization kit (*home*RNA) to capture transcriptomic signatures of disease and track their evolution over time in an out-of-clinic (remote) unsupervised use setting. We conducted a longitudinal observational case-control study in a cohort of individuals with acute COVID-19 infection (*n* = 39) and healthy controls (*n* = 5) (**Fig. 2A**). All healthy controls were recruited from the general population. For the control group, healthy adults with no history of respiratory symptoms or SARS-CoV-2 positivity within 14 days of eligibility screen were recruited from the general population. In the COVID-19+ group, both vaccinated and unvaccinated COVID-19+ adult participants with a positive SARS-CoV-2 nucleic acid amplification test within 7 days of eligibility screen were recruited through the COVID-19 Clinical Research Center. Participants who completed the full COVID-19 vaccination series at least two weeks prior to study enrollment were classified as vaccinated. Participants who received at least a single dose of COVID-19 vaccination but did not meet the two-weeks requirement above were classified as partially vaccinated. Participants who did not receive any doses of COVID-19 vaccination prior to or during their participant were classified as unvaccinated. All study participants provided informed consent. Each study participant collected blood samples using *home*RNA every other day (7 sampling timepoints), collected daily nasal swab samples (14 sampling timepoints) and completed daily symptom surveys (14 surveys) over a two-week period to track blood transcriptional response, viral load kinetics, and symptom progression respectively. The study was approved by the Fred Hutchinson Institutional Review Board [protocol approval number: FH10523]. This study was conducted at the Fred Hutchinson Cancer Center, Seattle WA. All sample collections were performed remotely by study participants. Participants were recruited to the study between January – September 2021.

### Respiratory specimen collection and viral load kinetics

COVID-19+ participants were asked to collect anterior nasal swab samples from both nostrils using a sterile polyurethane custom foam swab (Puritan Ref# 251805PFSC2ARROW). Respiratory swab specimens were stored dry (without universal transport medium), mailed back to the lab, and immediately transferred to -80°C storage until ready for pathogen analysis. To quantify viral load, total nucleic acid was extracted using Magna Pure 96 small total nucleic acid isolation kit (Roche Diagnostics). Isolated nucleic acid was screened for the presence of SARS-CoV-2 and multiple other respiratory viral and bacterial pathogens (**Table S5**) by TaqMan-based quantitative reverse transcription polymerase chain reaction (RT-qPCR) on the OpenArray platform (Thermo Fisher). TaqMan relative threshold values (Crt) were used to estimate viral load. A pathogen was classified as detected in a respiratory specimen when its Crt value was ≤ 28. To track viral load kinetics, respiratory specimens with time-matched *home*RNA blood samples were serially assayed until samples were negative for any respiratory pathogen in two consecutive samples.

### Participant reported outcomes and *home*RNA device use surveys

Daily symptom surveys were administered virtually via REDCap (*41*) in the COVID-19 case group. A total of 26 respiratory and non-respiratory symptom categories were presented in the survey (**Table S6**). Participants scored each symptom category based on severity (0 = none, 1 = mild, 2 = moderate, 3 = severe). COVID-19 vaccination manufacturer and dates were obtained to further classify COVID-19 cases into vaccinated and unvaccinated subgroups. Participants also completed a device use survey after each *home*RNA blood collection to assess device usability, kit integrity during transport, blood collection parameters, and pain levels experienced during *home*RNA blood collection.

### Gene expression analysis

#### RNA isolation, cleanup, and concentration

All procedures using commercial kits were performed according to the manufacturer’s recommended protocol unless otherwise noted. For PAXgene venipuncture samples, total RNA was extracted using the PAXgene Blood RNA kit. For *home*RNA-stabilized blood samples, total RNA was isolated using the Ribopure^™^ Blood RNA Isolation Kit. RNA concentrations were measured using the Take3 microvolume plates on the BioTek Cytation 5 multimode reader (Agilent technologies). RNA quality was measured on the Bioanalyzer 2100 (Agilent Technologies) using the RNA 6000 Nano kit (Agilent Technologies #5067-1511) for samples with concentrations ≥ 5 ng/µL. RNA samples with concentration < 5 ng/µL, RIN values were measured using the RNA 6000 Pico kit (Agilent Technologies #5067-1513). For nCounter gene expression analysis, RNA samples were column purified and concentrated using the Monarchâ RNA Cleanup Kit (NEB #T2030). Samples were stored at -80°C until ready for gene expression analysis.

#### nCounter analysis

For gene expression analysis, direct detection and digital counting of native RNA transcripts was performed on the nCounter Pro Analysis System (nanoString). For each participant sample, 50-100 ng of total RNA samples were hybridized to the nCounter Host Response Panel codeset (nanoString) containing both capture and molecular barcoded-reporter probes to target genes. 773 genes associated with the host response to infectious diseases and 12 candidate reference (housekeeping) genes were targeted. Target-probe hybrids were immobilized on a cartridge, aligned, and digitally counted on the nCounter Pro digital analyzer. Two Host Response codeset versions (Host Response v1.0 and Host Response v1.1) were used to generate the dataset. A panel standard containing identical counts of all target panel genes was run alongside participant samples for each of the two codeset versions to normalize counts between the two codeset.

### Statistical analyses

#### Differential gene expression analysis

We used generalized additive mixed models (GAMMs) to explore associations between gene expression and COVID-19 disease status over time, adjusting for age, sex, nCounter Host Response codeset versions (v1.0 and v1.1), and vaccination status (*12*). All fitted GAMMs included subject-specific random intercepts to account for potential intra-subject correlation and described longitudinal gene expression data using smoothed functions of time, defined as the number of days since first positive PCR test or onset of symptoms for COVID-19 positive participants. For healthy uninfected participants, an initial time point (Day 1) was defined as the median initial time point for COVID-19+ participants. We developed three GAMMs: i) *Model 1* included COVID-19 disease status as factor; ii) *Model 2* adjusted for vaccination but not COVID-19 disease status, and was fitted to data from COVID-19+ participants only; iii) *Model 3* was similar to *Model 1*, except that it adjusted for vaccination and COVID-19 disease status using a 4-level categorical variable with levels defined as (1) (COVID-19 negative, fully vaccinated), (2) (COVID-19 positive, unvaccinated), (3) (COVID-19 positive, partially vaccinated), and (4) (COVID-19 positive, fully vaccinated). P-values were adjusted for multiple comparisons by controlling the false discovery rate (FDR) using the Benjamini-Hochberg (BH) procedure (*42*). Adjusted p-values < 0.1 were used to identify differentially expressed transcripts. GAMM analyses were carried out using the gamm4 R package (*43*), setting the number of bases set to 5 in all analyses (*43*). Volcano plots was generated in R Statistical Software (v4.2.1, R Core Team 2022) (*44*). Functional enrichment was performed using graphical user interface ShinyGO (*45*).

### Time-course geneset analysis (TcGSA)

Dynamic genesets between various disease groups were identified using the TcGSA package available on CRAN (http://cran.r-project.org/web/packages/TcGSA/index.html) (*13*). The TcGSA analysis utilized mixed models to compute the likelihood ratios for queried genesets (*13*). To account for the heteroskedasticity of the nCounter gene expression data, regularized log (rlog) transformation of normalized nCounter gene expression counts was performed in DESeq2 (http://bioconductor.org/packages/DEseq2/) prior to TcGSA analysis (*46*) (**Fig. S5**). BloodGen3 (*14, 15*) geneset module was used to query dynamic genesets within our expression dataset. To ensure samples from individual participants are represented within each analyzed time-course group, sampling timepoints were used as the time variable in the analysis. Dynamic genesets between disease groups (COVID-19+ unvaccinated, COVID-19+ vaccinated, and healthy uninfected controls) were analyzed adjusting for age, sex, and codeset version using a linear time function. P-values were adjusted for multiple comparisons by controlling the FDR using the Benjamini-Hochberg (BH) procedure (*42*). Adjusted p-values < 0.1 were used to identify significant genesets. For TcGSA heatmap visualization, significant genesets were clustered using the squared Euclidean (Ward D2) distance and the number of gene expression clusters within each dynamic geneset was estimated using the default gap statistic parameters (K-max = 4; methods = “firstSEmax”) and 500 Monte Carlo bootstrap samples. Estimation of gene expression dynamics for each significant geneset obtained from the TcGSA likelihood ratio test were displayed in a heatmap to visualize global temporal trends. Individual genesets were visualized using the rlog expression values for each gene within the queried geneset and aligned to sampling timepoints 1 – 7 (T1-T7) and displayed as spaghetti plots with each solid blue line representing the median scaled gene expression for a given gene over all participants.

## Data Availability

All data are available in the main text or the supplementary materials. Gene expression data available upon request. All statistical analyses were conducted using R. Packages used to perform analyses are specified in the statistical section. Scripts developed to perform analyses are available upon request.

## List of Supplementary Materials

1. Detailed methods and materials

2. Supplemental figures S1-S4.

3. Supplemental tables S1-S6.

4. Data file S1. Normalized counts of nCounter gene expression dataset obtained in the study.

5. Data file S2. GAMM significant genes.

## Author contributions

We would like to thank Drs. Damien Chaussabel, Boris Hejblum, and Darawan Rinchai for project discussions. We would also like to thank the study participants for their participation in this research.

Conceptualization: FYL, ABT, AW

Methodology: FYL, SYK, AJH, OH, ABT, AW

Investigation: FYL, SYK, LMS, OH, ABT, AW

Visualization: FYL, SYK, HGL

Funding acquisition: ABT, AW

Project administration: KK, RLB, HGL, MSG

Supervision: LMS, MB, OH, ABT, AW

Writing – original draft: FYL, ABT, AW

Writing – review & editing: FYL, SYK, LMS, MB, JTS, OH, ABT, AW

