## Supplemental methods, figures, and tables for "Longitudinal home self-collection of capillary blood using *home*RNA correlates interferon and innate viral defense pathways with SARS-CoV-2 viral clearance"

**Supplemental Information**

**Detailed methods and materials**

***home*RNA blood collection**

Detailed characterization of the *home*RNA kit usage has been previously described (*11*). Briefly, participants were instructed to use the Tasso-SST blood collection device on the upper arm. General *home*RNA blood collection procedure included i) warming the collection site using a heat pack to facilitate blood flow, ii) cleaning the site with provided alcohol wipes, iii) adhering the Tasso-SST blood collection device to the cleaned area, iv) pressing a button to activate the lancet and initiate blood collection, v) removing the Tasso-SST device after five minutes or when the collection tube is full (whichever comes first), and vi) covering the puncture site with provided sterile bandage. Immediately after blood collection, participants were instructed to disconnect the Tasso-SST blood tube (**Fig. 1**) and reconnect it to the stabilizer tube to initiate mixing of the collected blood and stabilizer. The stabilized blood samples were transferred into a 50-mL conical tube containing a custom-design tube insert and mailed back to the University of Washington at ambient temperature using overnight courier services directly to a secure -20ºC freezer. Returned blood samples were transferred to a -80ºC freezer within 1-2 days post-receipt for storage until ready for RNA extraction.

**Comparison between PAXgene venipuncture versus *home*RNA self-collection RNA quality.**

For comparison between PAXgene venipuncture and *home*RNA self-collection methodologies for RNA yield and quality, healthy volunteers ($n$ = 20) were recruited from the general population (**Fig. S1A**). We collected a single PAXgene venipuncture sample from each participant on-site. Participants were then given a *home*RNA blood sampling kit and asked to self-collect and stabilize capillary blood samples at home on the same day. *home*RNA-stabilized blood samples were mailed back to the lab for RNA extraction and analysis.

***home*RNA blood sampling kit assembly and device fabrication.**

Detailed description of the *home*RNA kit and device design has been previously described (*11*). Briefly, the Tasso-SST blood collection device was purchased from Tasso, Inc. The RNA stabilizer reagent vial, adaptor, and cap were injection molded out of polycarbonate (PC: Makrolon 2407) by Protolabs, Inc (Maple Plain, MN). Prior to fabrication, all components of the stabilizer tube were sonicated in 70% ethanol (v/v) for 30 minutes and air dried. The adaptor was bonded onto the reagent vial using Dymax MDÒ UV-curable medical grade glue (#1450-M-UR-SC). Bonded parts were UV-cured for 12 minutes at 30ºC using a Form Cure UV resin-curing chamber (Formlabs). The fabricated stabilizer vial was filled with 1.3 mL of RNA-stabilizing solution (RNAlater™), capped, and packaged in an impulse-sealed Tyvek pouch. Each *home*RNA blood collection kit was assembled with a Tasso-SST blood collection device, a stabilizer tube, instructions for use, and all other kit components required for the participant to perform blood collection as previously described (*11*).

**SARS-CoV-2 sequencing**

Sequencing was attempted for a single SARS-CoV-2 positive respiratory sample with the lowest Crt value (highest viral load) from each COVID-19+ participant. Nucleic acid was extracted using the Magna Pure 96 small total nucleic acid isolation kit (Roche Diagnostics) and sequencing libraries prepared using the COVIDSeq kit (Illumina). Artic V4 primers were used (https://community.artic.network/t/sars-cov-2-version-4-scheme-release/312). Viral genomes were sequenced using the NextSeq2000 P200 kit (Illumina), and the SARS-CoV-2 reference genome (Wuhan/Hu-1/2019; Genbank accession MN908947) was used to assemble consensus genomes using a modified iVar pipeline. Nextstrain augur software was used to align viral sequences and construct phylogenetic trees for variant determination.

**nCounter data quality control and normalization.** nCounter RCC files of all samples were pre-processed on the nSolver™ software (nanoString) to obtain normalized expression counts. Quality control (QC) measures were applied to all participant samples. Imaging QC was assessed by measuring the percent field of view (FOV) successfully scanned within each sample. Samples with scanned FOV < 75% were flagged for removal. Binding density QC was assessed by measuring the density of the reporter probe on the cartridge surface within each sample. Samples with binding density outside of 0.1 - 2.25 spots/square micron were flagged for removal. The correlations between the observed counts for all six positive ERCC control probes (Positives A – F) and their spike-in synthetic nucleic acids (0.125 fM – 128 fM) were used to determine a sample's positive control linearity. Samples with positive control linearity of < 0.95 were flagged for removal. Gene expression count normalization was performed on all samples that passed the QC metrics above. For normalization, all samples were subjected to i) positive control normalization, ii) codeset content normalization, and iii) panel standard normalization. Selection of reference genes was performed using ROSALIND® (https://rosalind.bio/) based on nanoString recommendations. Nine reference genes (*ABCF1, GUSB, HRPT1, MRPS7, NMT1, NRDE2, PGK1, SDHA, TBP*) with stable expression within the dataset as determined by geNorm were used in codeset content normalization .

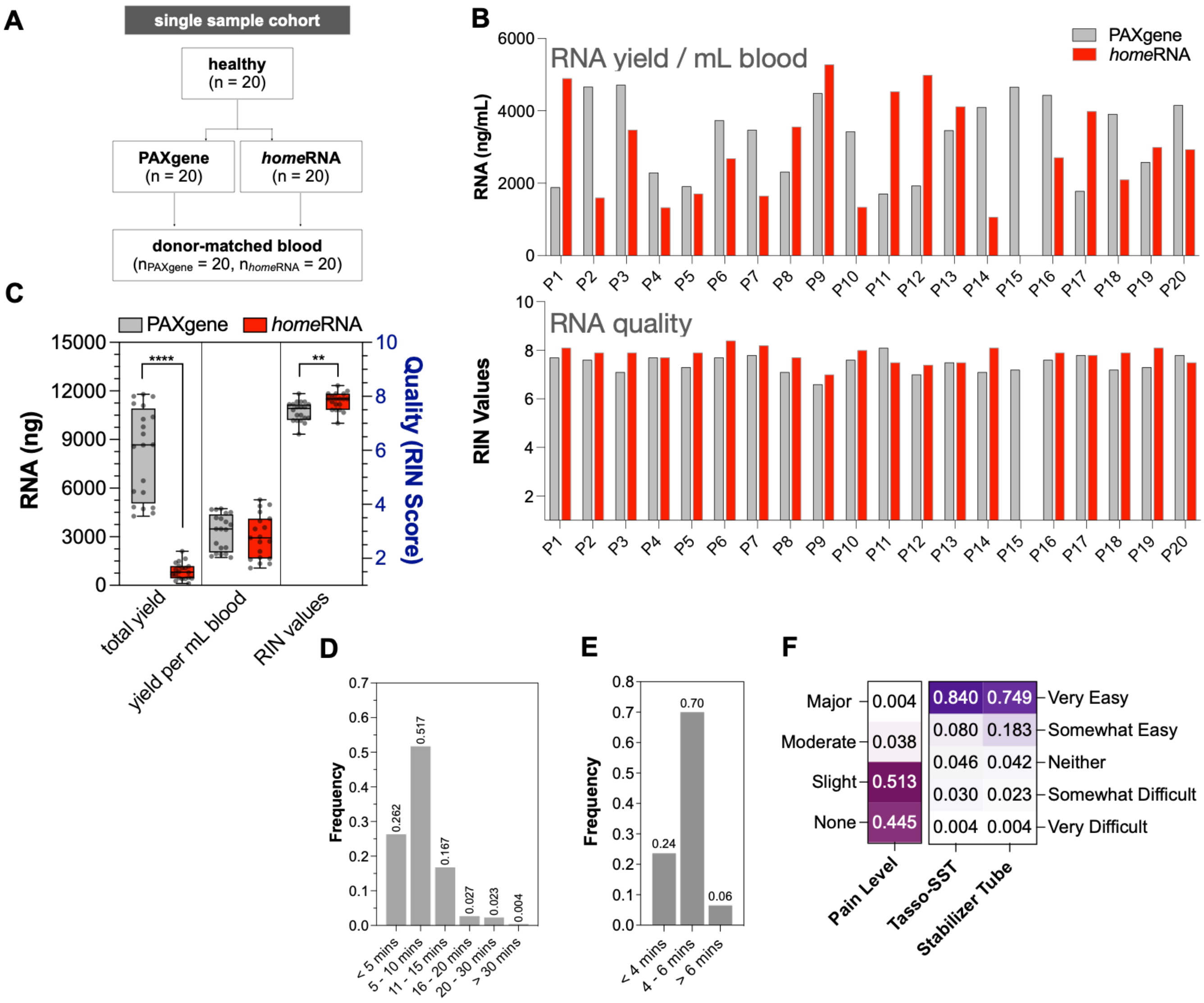


**Figure S1. *home*RNA capillary blood provided comparable RNA yield and quality compared to donor-matched PAXgene venous blood. A**) Flowchart of study cohort. **B**) RNA yield (per mL blood volume) and RNA quality (RIN) comparing *home*RNA (red bars) and PAXgene (gray bars) samples in individual participants. **C**) Box and whisker plot showing total RNA yield, RNA yield per mL blood, and RIN values comparing *home*RNA and PAXgene samples. **D**) *home*RNA kit usage time. **E**) Tasso-SST blood collection time. **F**) Reported pain level and device usability (stratified by blood collection and stabilization steps).

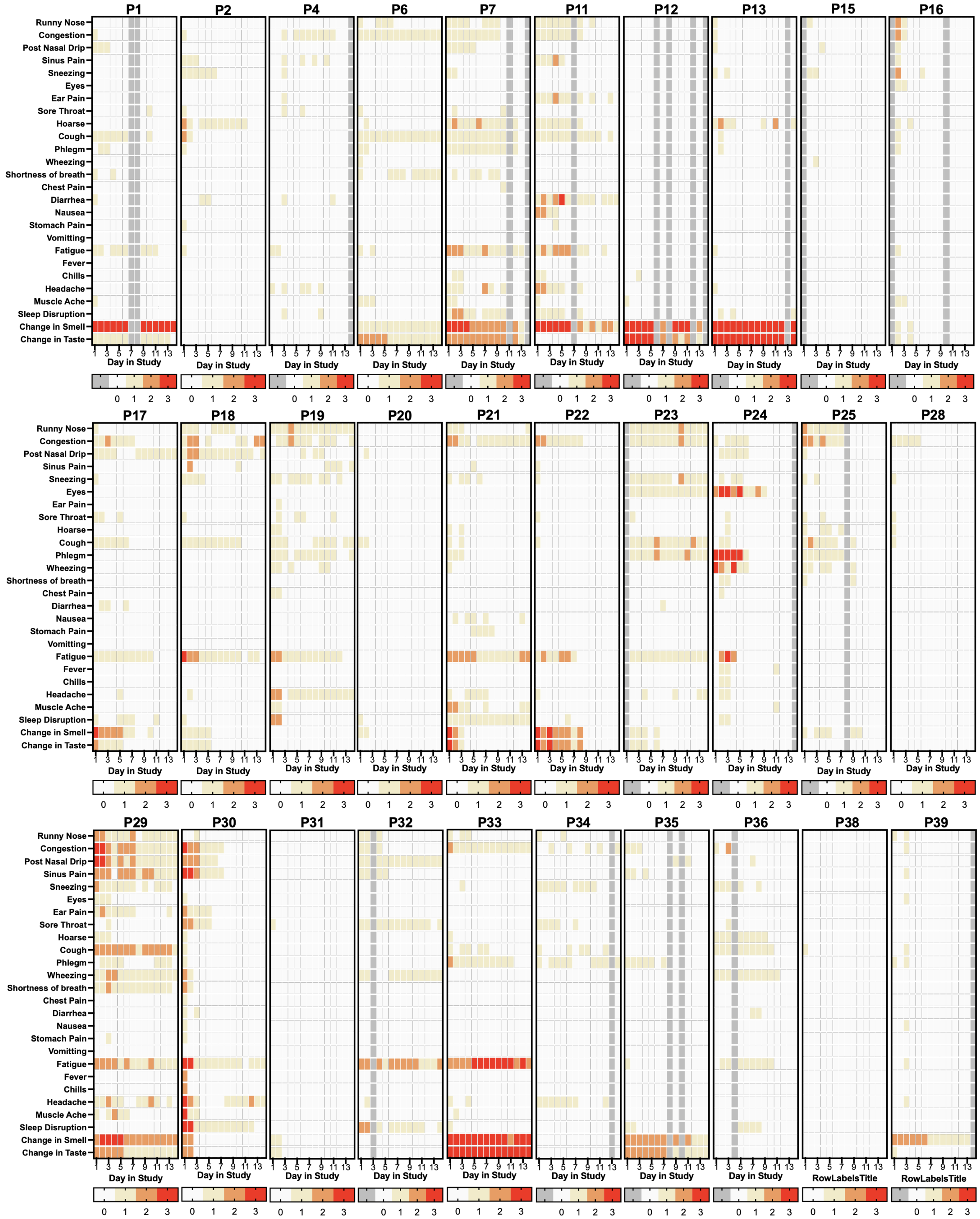


**Figure S2. All isolated RNA obtained from decentralized blood collection using *home*RNA passed QC parameters. A**) % field-view imaged, **B)** positive control linearity (%), **C**) binding density scores, and **D**) limit of detection for all assayed samples. Each floating bar plot represents a single assay consisting of twelve total samples. All samples were processed across four total run batches.

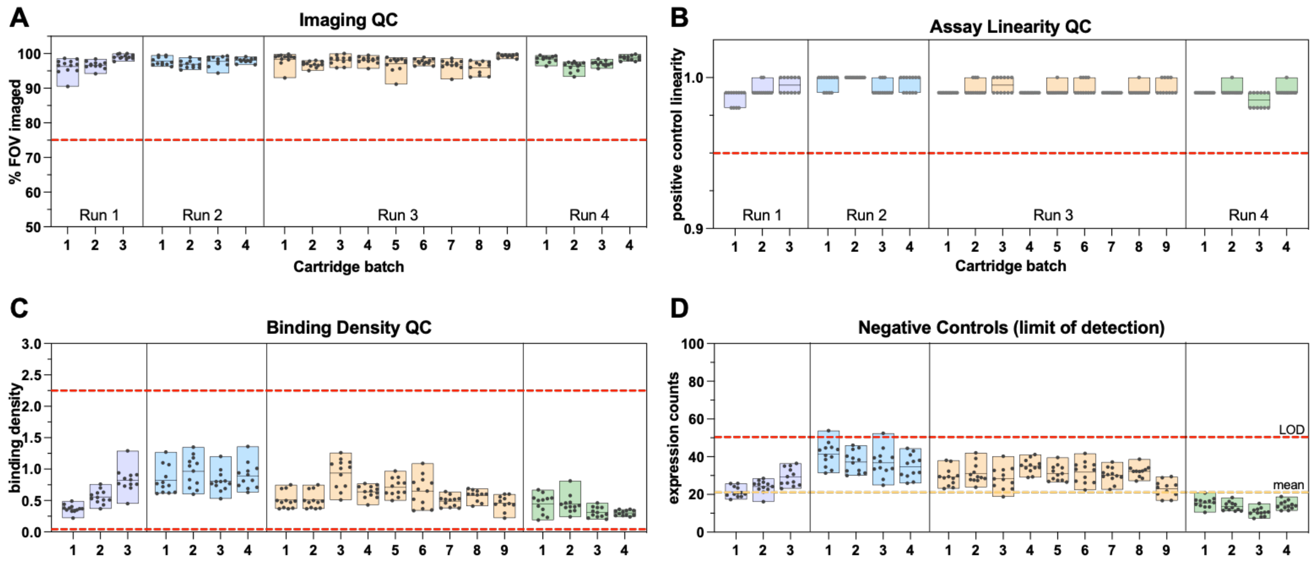


**Figure S3. Symptom burden in individual participants.** Symptom severity is scored from 0-3 (0= no symptoms; 3 =severe symptoms). Gray cells denote missing survey response.

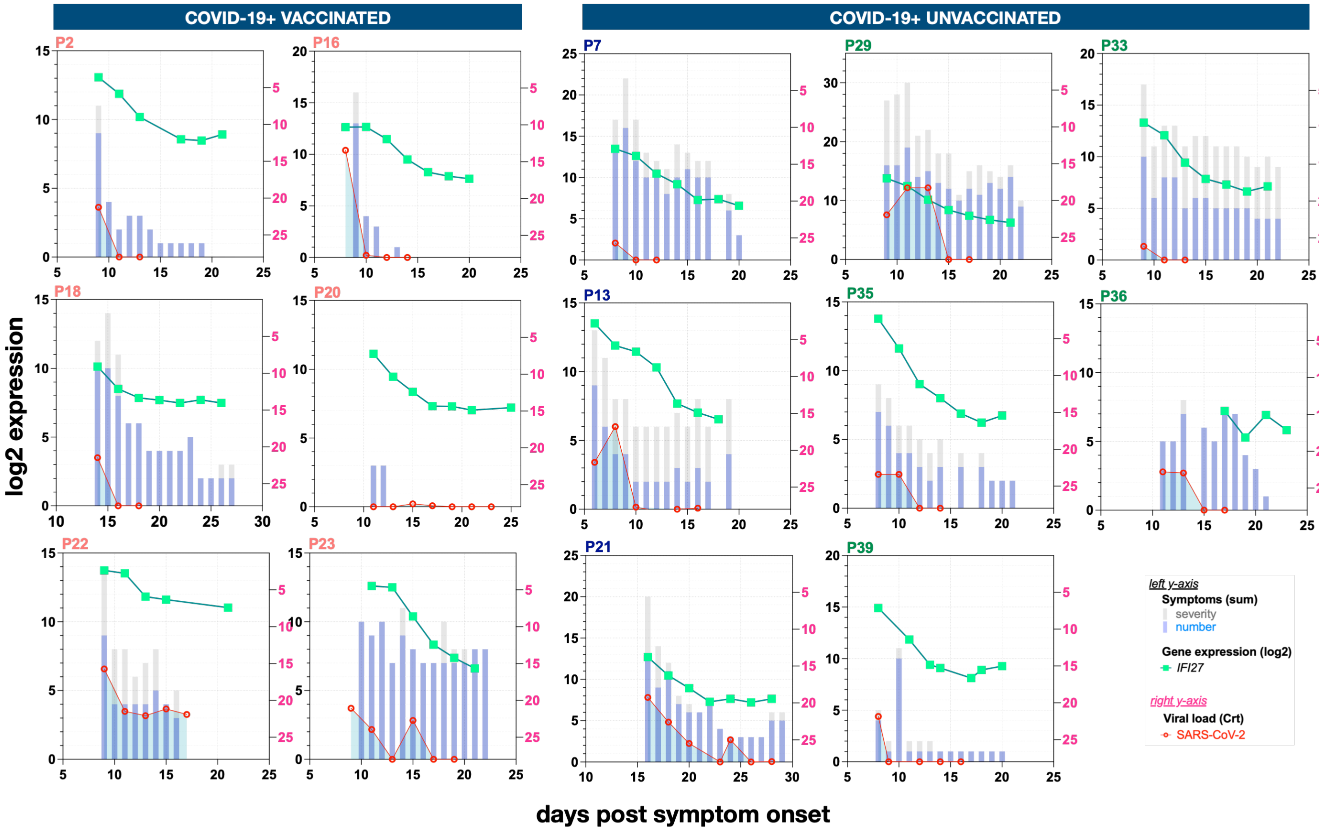


**Figure S4. Kinetics of *IFI27* gene expression, viral load, and symptoms in individual participants.** Partially and fully vaccinated participants are labeled in blue and green respectively. Total symptom number (blue columns), total symptom severity (gray columns), and *IFI27* gene expression kinetics are plotted on the left *y-axis*. SARS-CoV-2 viral load is plotted on the right *y-axis*. *IFI27* expression kinetics of all healthy uninfected controls are displayed on the far-right plot.

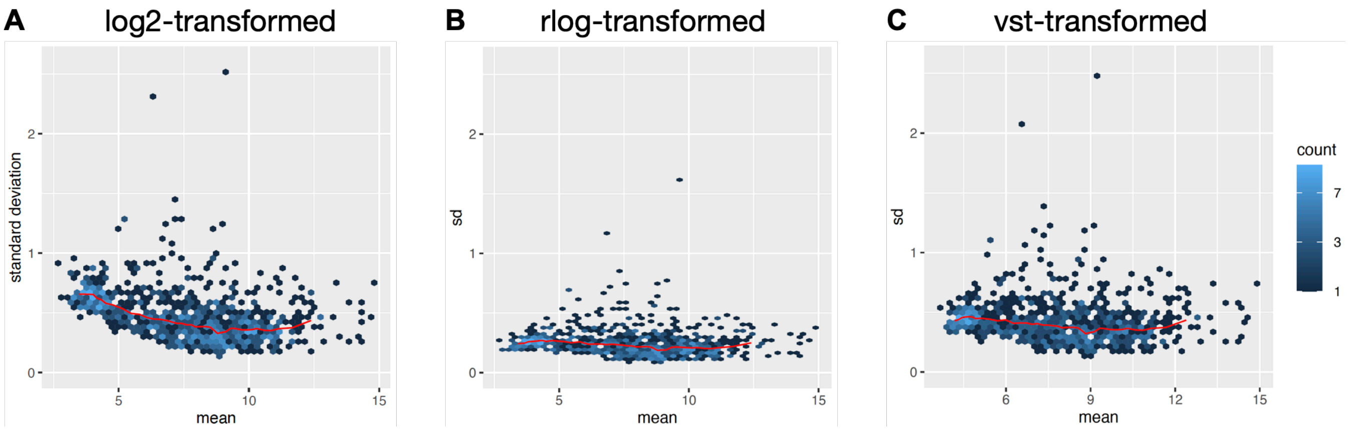


**Figure S5. Dependence of variance on the mean across transformation types.** Scatterplot of mean-standard deviation (MSD plot) of **A)** log2 transformation (log2), **B)** regularized log (rlog) transformation (rlog), and **C)** variance-stabilizing (vst) transformation. Red line depicts the running median estimator with 10% window-width.

### SUPPLEMENTAL TABLES

#### Table S1. Summary of scheduled and returned blood and nasal swab specimens.

| Disease Status | Healthy | COVID-19 | Total | % total |
|---|---|---|---|---|
| Participants | | | | |
| **Enrolled** | 5 | 39 | 44 | |
| **Withdrew** | 0 | 2 | 2 | 4.5% |
| **Excluded**<br>- incomplete collection<br>- low RNA yield | <br>0<br>0 | <br>6<br>1 | 7 | 15.9% |
| **Analyzed** | 5 | 30 | 35 | 79.5% |
| *home*RNA blood samples (*n* = 36) | | | | |
| **Scheduled** | 35 | 217 | 252 | |
| **Received** | 34 | 213 | 247 | 98.0% |
| **Analyzed** | 31 | 201 | 232 | 92.1% |
| Respiratory swabs (*n* = 30) | | | | |
| **Scheduled** | none | 420 | 420 | |
| **Received** | none | 413 | 413 | 98.3% |
| **Analyzed** | none | 168 | 168 | 40.0% |

#### Table S2. *home*RNA collection parameters

| Blood Level | Volume (µL) | Sample number | Frequency | Returned | Analyzed | Not Analyzed | Yield (ng); Median (IQR) |
|---|---|---|---|---|---|---|---|

| | | | | | | | |
|---|---|---|---|---|---|---|---|
| No blood | 0 | 3 | 0.0119048 | | | | |
| Level 1 | 100 | 42 | 0.1666667 | 42 | 33 | 9 | 762.9 (448.4-1262) |
| Level 2 | 200 | 31 | 0.1230159 | 31 | 30 | 1 | 872.1 (554.7-1153) |
| Level 3 | 300 | 53 | 0.2103175 | 52 | 49 | 3 | 1049 (729.8-1572) |
| Level 4 | 400 | 115 | 0.4563492 | 115 | 114 | 1 | 1541 (852.2-2370) |
| Not reported | unknown | 8 | 0.031746 | 7 | 6 | 1 | |
| **Total Samples** | | 252 | | 247 | 232 | 15 | |

**Table S3. SARS-CoV-2 variants in COVID-19+ unvaccinated and breakthrough infections**

| SARS-CoV-2 Variant | Unvaccinated N (%) | Vaccinated N (%) |
|---|---|---|
| 20A | 1 (7) | 0 (0) |
| 20B | 2 (14) | 1 (6) |
| Alpha | 6 (43) | 1 (6) |
| Epsilon | 4 (29) | 2 (13) |
| Delta | 0 (0) | 9 (56) |
| Unknown | 1 (7) | 3 (19) |

**Table S4. Parameters of fitted GAMM models**

| GAMM | Covariates | Vaccination Level | Samples analyzed | Contrast groups | Smoothed functions |
|---|---|---|---|---|---|
| Model 1 | age<br>sex<br>codeset<br>disease | | all participants | covid19:healthy | s(days):healthy<br>s(days):covid19 |
| Model 2 | age<br>sex<br>codeset<br>vaccination | Unvaccinated<br>Vaccinated (Partial)<br>Vaccinated (Full) | covid-19+ participants only | covid19 vacc:unvacc<br>covid19 partial:unvacc | s(days):unvacc<br>s(days):vacc(partial)<br>s(days):vacc(full) |
| Model 3 | age<br>sex<br>codeset<br>vaccination | Unvaccinated<br>Vaccinated (Partial)<br>Vaccinated (Full)<br>Healthy | all participants | covid19 unvacc:healthy<br>covid19 vacc:healthy<br>covid19 partial:healthy | s(days):healthy<br>s(days):unvacc<br>s(days):vacc(partial)<br>s(days):vacc(full) |

**Table S5. List of pathogens screened in respiratory specimen.**

| Pathogen Category | Strains |
|---|---|
| Influenza | Pan Flu A, Flu A (H3N2), Flu A (H1N1), Pan Flu B, Pan Flu C |
| Parainfluenza | Human Parainfluenza 1 - 4 |
| Enterovirus | Pan enterovirus and Enterovirus D68 |

| | |
|---|---|
| Rhinovirus | Rhinovirus 1 and 2 |
| Adenovirus | Adenovirus 1 and 2 |
| non-SARS coronavirus | HKU1, NL63, 229E, OC43 |
| SARS coronavirus | SARS-CoV-2 |
| Respiratory synctial virus | RSV A and RSV B |
| Metapneumovirus | hMPV |
| Bacterial pneumonia | *Streptococcus pneumoniae, Mycoplasma pneumoniae, Chlamydia pneumoniae* |

**Table S6. List of symptoms obtained from patient surveys.**

| Nose | | Eyes / Ears | | Throat | | Chest | |
|---|---|---|---|---|---|---|---|
| Runny nose | A1 | Watery eyes | B1 | Sore throat | D1 | Cough | E1 |
| Congestion | A2 | Ear Pain | C1 | Hoarse | D2 | Phlegm | E2 |
| Post nasal drip | A3 | | | | | Wheezing | E3 |
| Sinus pain | A4 | | | | | Shortness of breath | E4 |
| Sneezing | A5 | | | | | | |

| Gastrointestinal | | General | | Sleep / Sensory | |
|---|---|---|---|---|---|
| Diarrhea | F1 | Fatigue | G1 | Sleep disruption | H1 |
| Nausea | F2 | Fever | G2 | Change in smell | S1 |
| Stomach pain | F3 | Chills | G3 | Change in taste | S2 |
| Vomiting | F4 | Headache | G4 | | |
| | | Muscle ache | G5 | | |
